# Genetic and phenotypic heterogeneity in early neurodevelopmental traits in the Norwegian Mother, Father and Child Cohort Study

**DOI:** 10.1101/2023.09.20.23295829

**Authors:** Laura Hegemann, Elizabeth C. Corfield, Adrian Dahl Askelund, Andrea G. Allegrini, Ragna Bugge Askeland, Angelica Ronald, Helga Ask, Beate St Pourcain, Ole A. Andreassen, Laurie J. Hannigan, Alexandra Havdahl

**Affiliations:** Nic Waals Institute, Lovisenberg Diaconal Hospital, Oslo, Norway; Department of Psychology, University of Oslo, Oslo, Norway; Centre for Genetic Epidemiology and Mental Health, Norwegian Institute of Public Health, Oslo, Norway; Department of Clinical, Educational & Health Psychology, Division of Psychology & Language Sciences, Faculty of Brain Sciences, University College London, London, United Kingdom; Social, Genetic and Developmental Psychiatry Centre, Institute of Psychiatry, Psychology & Neuroscience, King’s College London, London, United Kingdom; NORMENT Centre, Institute of Clinical Medicine, University of Oslo and Division of Mental Health and Addiction, Oslo University Hospital, Oslo, Norway; School of Psychology, Faculty of Health and Medical Sciences, University of Surrey, Guildford, UK; PROMENTA Research Centre, Department of Psychology, University of Oslo, Oslo, Norway; Language and Genetics Department, Max Planck Institute for Psycholinguistics, Nijmegen, the Netherlands; MRC Integrative Epidemiology Unit (IEU), University of Bristol, Bristol, United Kingdom; Donders Institute for Brain, Cognition and Behaviour, Radboud University, Nijmegen, the Netherlands

## Abstract

Different neurodevelopmental conditions such as autism and ADHD frequently co-occur. Overlapping traits and shared genetic liability are potential explanations. We examine this using data from the population-based Norwegian Mother, Father, and Child Cohort study (MoBa), leveraging item-level data to explore the phenotypic factor structure and genetic architecture underlying neurodevelopmental traits at age 3 years (N = 41 708 – 58 630). We identified 11 latent factors at the phenotypic level using maternal reports on 76 items assessing children’s motor skills, language, social functioning, communication, attention, activity regulation, and flexibility of behaviors and interests. These factors showed associations with diagnoses of neurodevelopmental conditions and most shared genetic liabilities with autism, ADHD, and/or schizophrenia. Item-level GWAS revealed trait-specific genetic correlations with autism (item *r*_g_ range = -0.27 – 0.78), ADHD (item *r*_g_ range = -0.40 – 1), and/or schizophrenia (item *r*_g_ range = -0.24 – 0.34). Based on patterns of item-level genetic covariance and genomic factor analyses, we find little evidence of common genetic liability across all neurodevelopmental traits. These results more so support genetic factors across more specific areas of neurodevelopment, some of which, such as prosocial behavior overlap with factors found in the phenotypic analyses. Other areas such as motor development seemed to have more heterogenous etiology, with indicators in this domain showing a less consistent pattern of genetic correlations with each other. Overall, these exploratory findings emphasize the etiological complexity of neurodevelopmental traits at this early age. In particular, diverse associations with neurodevelopmental conditions and genetic heterogeneity could inform follow-up work to identify shared and differentiating factors in the early manifestations of neurodevelopmental traits, which in turn could have implications for clinical screening tools and programs.

## Introduction

Recent versions of international diagnostic classification systems have introduced an umbrella category of neurodevelopmental conditions [1,2]. Conditions classified in this category typically manifest from childhood and are characterized by divergent trajectories of development. Generally, they are diagnosed based on significant difficulties in developmental skills in areas such as language, social abilities, learning, or motor activity. Conditions such as attention-deficit hyperactivity disorder (ADHD), autism spectrum conditions (autism), intellectual disabilities, specific learning disabilities, developmental coordination disorder and tic conditions are now classified together; but have previously been conceptualized as independent and sometimes mutually exclusive conditions. For example, under DSM-IV, autism was an exclusion criterion for ADHD preventing their co-diagnosis. However, there is increasing evidence that conditions in this category share many characteristics, such as high heritability [3–5]; heterogeneous clinical presentation with a wide range of support needs [6,7]; and marked sex differences, with higher prevalence in males [8–10].

In addition to shared characteristics, neurodevelopmental conditions frequently co-occur [11,12], as do their symptoms at sub-diagnostic threshold levels [13,14]. While the etiology of this co-occurrence is not well understood, some observations have implicated shared genetic liability between neurodevelopmental conditions. For example, siblings of an individual with one neurodevelopmental condition often have increased likelihood for several neurodevelopmental conditions [15,16]. Additionally, unidentified latent genetic factors [17] as well as identified common [18–21] and rare genetic variants [22–24] are shared amongst many clinically distinct neurodevelopmental conditions. Overlapping symptomatology and a lack of clear diagnostic boundaries have led to continuing revisions of the classifications of neurodevelopmental conditions [25–28].

Investigating the genetic and nosological bases for co-occurring neurodevelopmental conditions requires detailed data on their traits. Population-based registries are typically limited to diagnostic (yes/no) outcomes. Clinical cohorts, which may have more detailed data, are generally smaller and commonly ascertain individuals based on a single condition. Thus, meaningful analyses of common genetic variants and shared etiology across areas of development and specific traits are difficult. Data collected in population-based cohorts, typically have more breadth and depth of information that can help explore shared etiology of neurodevelopmental traits, but relatively fewer individuals with neurodevelopmental conditions. Still, relevant traits, capturing individual differences in language and motor development, attention, hyperactivity, social behavior, and repetitive, restricted behaviors and interests can be observed in all children. These traits are likely influenced by some of the same underlying genetic liabilities as neurodevelopmental conditions [29–34]. The prospective nature of population-based birth cohorts means these traits can be studied early – prior to or around the age at which neurodevelopmental diagnoses are most commonly made [35–37]. Exploring the relationships between neurodevelopmental traits early in life, investigating their genetic liabilities, and exploring links to neurodevelopmental conditions can give new insights about etiological mechanisms underlying the development and differentiation of such conditions.

In the present study, we leveraged detailed information about multiple traits related to different neurodevelopmental conditions. We investigated the phenotypic factor structure and genetic architecture underlying early (age 3 years) neurodevelopmental traits in a large population-based birth cohort. We additionally investigated, associations of these early signs with later neurodevelopmental conditions at both the phenotypic and genotypic level.

## Methods

### Measures and sample

#### Sample

The Norwegian Mother, Father and Child Cohort Study (MoBa) is a population-based pregnancy cohort study conducted by the Norwegian Institute of Public Health. [38,39] Participants were recruited from all over Norway from 1999-2008. The women consented to participation in 41% of the pregnancies. Blood samples were obtained from both parents during pregnancy and from mothers and children (umbilical cord) at birth. The cohort includes approximately 114 500 children, 95 200 mothers and 75 200 fathers. The current study is based on version 12 of the quality-assured data files released for research in January 2019. The establishment of MoBa and initial data collection was based on a license from the Norwegian Data Protection Agency and approval from The Regional Committees for Medical and Health Research Ethics. The MoBa cohort is currently regulated by the Norwegian Health Registry Act. The current study was approved by The Regional Committees for Medical and Health Research Ethics (2016/1702).

The present study was conducted on a subset of the cohort (n = 58 630) who had information available from the 36-month questionnaire. The children were an average of 3.1 years (SD = 0.18) old when mothers completed the questionnaire. The sample had 1.04:1 male: female ratio. Genetic analyses were conducted using a further quality controlled genotyped subset of the cohort (n = 42 934). For more information on genotyping of the MoBa sample and for the family-based quality control pipeline used to prepare these data for analysis, see Corfield et al. [40]

#### Measures for neurodevelopmental traits

We included items from all maternal report scales related to neurodevelopment in the 36-month questionnaire that asked about children’s observable behavior (as opposed to maternal concerns). This included items from the Social Communication Questionnaire (SCQ) [41], Ages and Stages Questionnaire (ASQ) [42], Non-Verbal Communication Checklist (NVCC)[43], Modified Checklist for Autism in Toddlers (M-CHAT) [44], Early Screening for Autistic Traits Questionnaire (ESAT) [45], the attention and hyperactivity questions from the Child Behavior Checklist (CBCL) [46], the prosocial behaviors subscale of the Strength and Difficulties Questionnaire (SDQ) [47] as well as several MoBa-specific questions. All items included had either dichotomous (e.g., yes/no) or trichotomous (e.g., not true/sometimes true/often true) response categories. Items were reverse coded where needed so that higher values reflected greater endorsement of the trait.

#### Measures for diagnostic and clinically relevant outcomes

Diagnostic data was ascertained from the Norwegian Patient Register (NPR) between 2008 and June 2021 based on ICD-10 criteria using MoBa *phenotools* [48]. Diagnostic groups were defined for receiving a diagnostic code at least one time for ADHD (F90), autism (F84.0, F84.1, F84.5, F84.8, and F84.9), intellectual disability and general developmental delay (F7 and F83), specific conditions of speech and language (F80, F98.5, F98.6), specific conditions of scholastic skills (F81), specific conditions of motor function (F82), and tic conditions (F95). The percent of the diagnostic group with NPR data available by age 3 who had received a diagnosis before age 4 varied by diagnosis from 0% (specific conditions of scholastic skills) to 26% (specific conditions of motor function).

Further information on the scales, the items used in the factor models, and diagnostic and clinically relevant outcomes are available in the supplementary methods and Supplementary Tables S1-3.

#### Polygenic scores

Polygenic scores (PGS) were estimated with the software PRSice2 [49] based on summary statistics from the most recent Psychiatric Genomic Consortium GWAS for ADHD [18], autism [19], and schizophrenia [50]. ADHD and autism were included as they are neurodevelopmental conditions with well powered and publicly available GWAS summary statistics. Schizophrenia was included given neurodevelopmental aspects to its development [51–53]. Scores were regressed on the first 10 genomic principal components (PCs) and genotype batch. The first principal component of 11 scores, constructed based on p-value thresholds between 5 × 10^-8^ and 1, was used for the subsequent analyses. This approach controls for type one error rate arising from optimization of pruning and thresholding while still maintaining prediction performance [54].

### Analyses

An overview of the analyses performed as well as thresholds for item inclusion in each analytic step are presented in Figure 1. Lenient thresholds for item selection were chosen to maximize the number of traits across different areas of development. Analytical code can be found at https://github.com/psychgen/neurodevelopment_traits_structure

**Figure 1:**
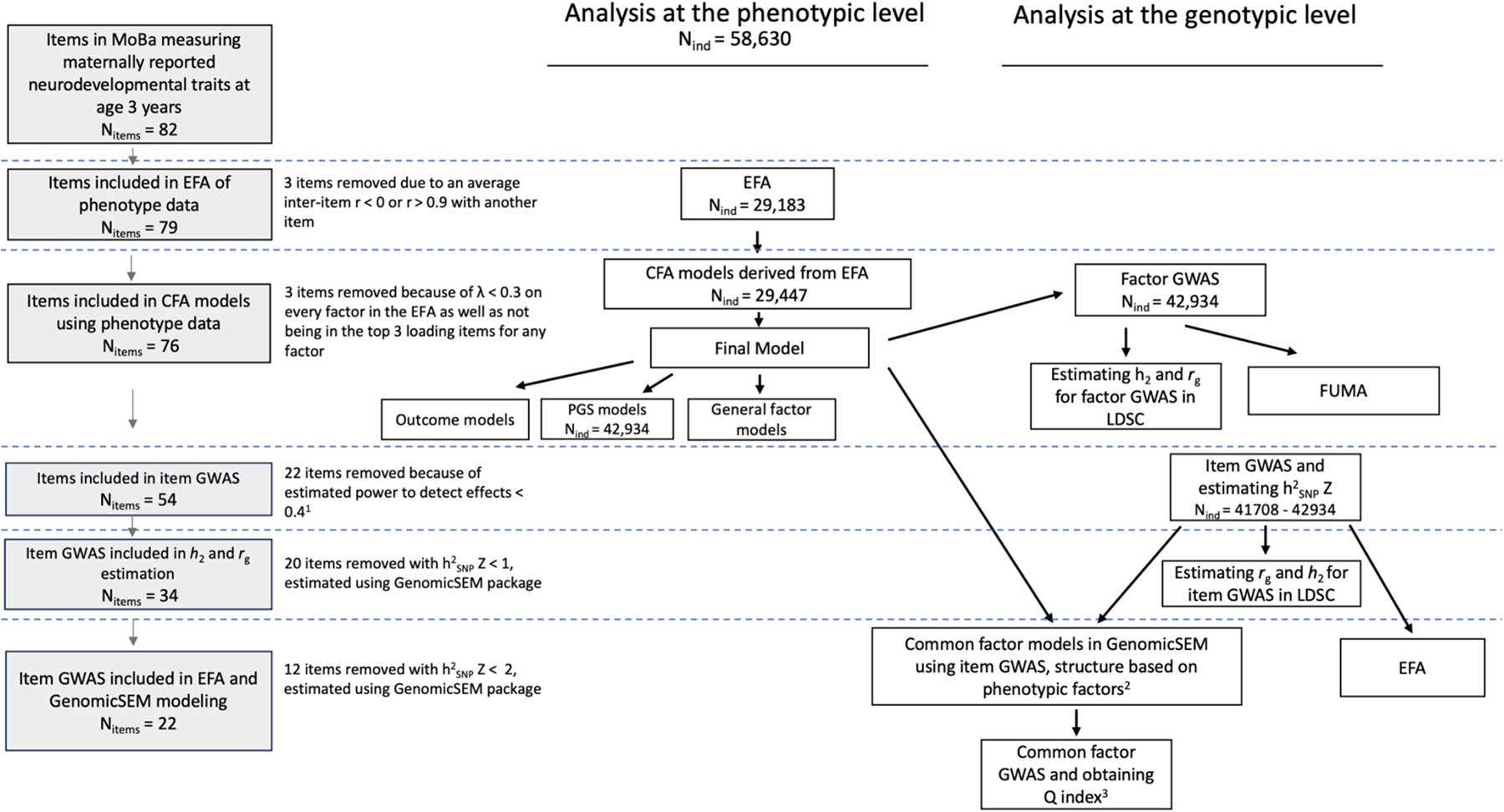
Outline of study design and main analyses at the phenotypic and genotypic level. Grey boxes outline the steps where questionnaire items were removed with the exclusion thresholds listed to the right. Boxes indicate an analysis with the arrows denoting analyses which are based on (i.e., factor structure) or used results (i.e., summary statistics) from a previous analysis. Analyses conducted at the phenotypic level with no sample size listed were conducted in the full sample (N = 58,630). Half-samples for the EFA/CFA conducted in the phenotypic level were randomly selected halves of the full sample. Estimating r_g_ refers to estimation of genetic correlations of the items/factors with neurodevelopmental conditions. ^1^ With the assumptions of an OR of 1.2, MAF of 0.01, and alpha of 0.01 in a logistic model with additive genetic effects. ^2^ Only common factor models with 3+ items run. ^3^ Common factor GWAS only run on models with good fits and significant factor loadings.

#### Exploratory and confirmatory factor analyses

Exploratory factor analysis (EFA) was performed in one randomly selected half of the full sample (n = 29 183). Confirmatory factor analyses (CFA) were run in the other half of the full sample (n = 29 447) for possible viable models derived from the EFA. Using standard fit indices (CFI, TLI, RMSEA) the best fitting model out of these possible models was used as the final model for all downstream analyses. In the full sample, to assess a unidimensional factor in addition to domain-specific factors, both bifactor and higher-order models were run alongside the final selected correlated factor model. To address potential sex differences in the measurement of these factors, we conducted measurement invariance testing in the full sample. A multi-group CFA (MG-CFA) of the correlated factor model by sex (N_males_ = 29 955, N_females_ = 28 589) was used to test for configural invariance and invariance of thresholds and loadings [55]. See supplementary methods for further details on the factor analyses, criteria for model selection, and measurement invariance testing.

#### Measurement models with neurodevelopmental diagnoses, clinically relevant outcomes, and polygenic scores

The factor associations with later outcomes served two purposes of 1) validation and further characterization of the factors; and 2) insight into how specific areas of development at age 3 are related to receiving a particular neurodevelopmental condition diagnosis. A correlated factor and a higher-order general factor model were run specifying the factors to predict neurodevelopmental diagnoses and other clinically relevant outcomes. In the higher-order model, general and specific factors were specified to predict outcomes separately in two models. Models were run in a multi-group SEM framework, grouped by sex with both regression effects and model parameters estimated for each sex separately. In the correlated factor models, both univariate models with the factors predicting the outcomes individually and multivariate models with factors predicting the outcome simultaneously were run. Due to collinearity concerns in the multivariate models arising from groups of highly correlated factors, the magnitude of the factors’ effects within those groups were constrained to be equal in the correlated factor model. Measurement models including PGS as explanatory variables for factors were run in the correlated factors and higher-order model.

#### Factor analyses software

EFA analyses were all run using the weighted least square mean and variance adjusted (WLSMV) estimation method and with a geomin oblique rotation applied in the Mplus statistical software (Muthén & Muthén, 2011). All CFA and measurement invariance models were run using the *lavaan* (v0.6-14) and *semTools* (v0.5-6) packages in R with the WLSMV estimation method [56,57]. Missing data was handled using pairwise deletion for both the EFA and CFA, as it is the default in Mplus for categorical data.

#### Genome-wide association studies

Genome-wide association studies (GWAS) were run on each individual item (item GWAS) for which power calculations indicated sufficient statistical power, and on factor scores estimated for each factor (factor GWAS). Factor scores were estimated using parameters for each sex from the correlated factor model multi-group CFA using the Empirical Bayes Model approach, the *lavaan* default method for categorical indicators. All GWAS included sex, genotype batch, and the first 10 PCs as covariates. Additional sex specific GWAS were run as sensitivity analyses for the factors. GWAS were run using version 3.1 of the REGENIE software, a computationally efficient linear mixed model method of conducting multi-trait GWAS in large samples using a two-step machine-learning paradigm. REGENIE can handle relatedness in the sample and correct for unbalanced case–control phenotypes in binary phenotypes [58]. For all factor and feasible item GWAS, SNP-based heritability (h^2^_SNP_) and genetic correlations (*r*_g_) with ADHD [18], autism [19], and schizophrenia [50] were estimated using linkage disequilibrium score regression (LDSC) [59]. Estimated h^2^_SNP_ for the item GWAS was on the liability scale. Functional mapping and annotation of the factor GWAS results were performed with FUMA (v1.5.3) [60]. Further information on sample sizes, prevalence estimates for LDSC, and power estimates used for the above analyses are listed in the supplementary methods and Supplementary Table S4.

#### Genomic factor modeling and specificity of SNP effects

Genomic factor modeling used selected item GWAS. A lenient item GWAS inclusion threshold (Figure 1) meant that power was borderline for genomic factor modeling, so an EFA was conducted on the estimated smoothed genetic correlation matrix of all chromosomes, as opposed to even/odd split, and no further downstream analyses (e.g., CFA) were conducted based on the results. Version 4.1.2 of the R package *stats* [61] was used to run the EFA and a promax rotation was applied. Common factor models based on factors from the phenotypic models that had at least three items meeting the item GWAS power threshold were run. For those with good fits and significant factor loadings a common factor GWAS was run estimating SNP and Q_SNP_ effects. Q_SNP_ being a measure of how well the association of the SNP and the individual trait is accounted for by the factor [21,62]. All confirmatory genomic factor modeling and GWAS were conducted using diagonally weighted least squares (DWLS) estimation in version 0.0.5 of the *GenomicSEM* R package [62].

## Results

### Phenotypic factor structure underlying early neurodevelopmental traits

Results of the EFA (Supplementary Tables S5-19) and CFA models indicated high dimensionality underlying early neurodevelopmental traits. Procedures to determine the optimal number of factors to retain indicated between 1-15 factors (Supplementary Figure S1) and fit indices from the EFA showed models with more than 9 factors met good fit criteria (Supplementary Table S5). Balancing these results with interpretability of the factors, 3 models (9,10 and 11-factor models) were selected to be run as confirmatory factor models in the other half of the sample. The 11-factor showed the best fit for complexity-penalized fit indices out of the three in both the EFA and CFA (Supplementary Tables S5, S20). A few poorly endorsed items had estimated loadings slightly over 1, constraining these values did not lead to a significant decrease in model fit. The 11-factor model was selected to be used in the downstream analyses.

**Figure 2:**
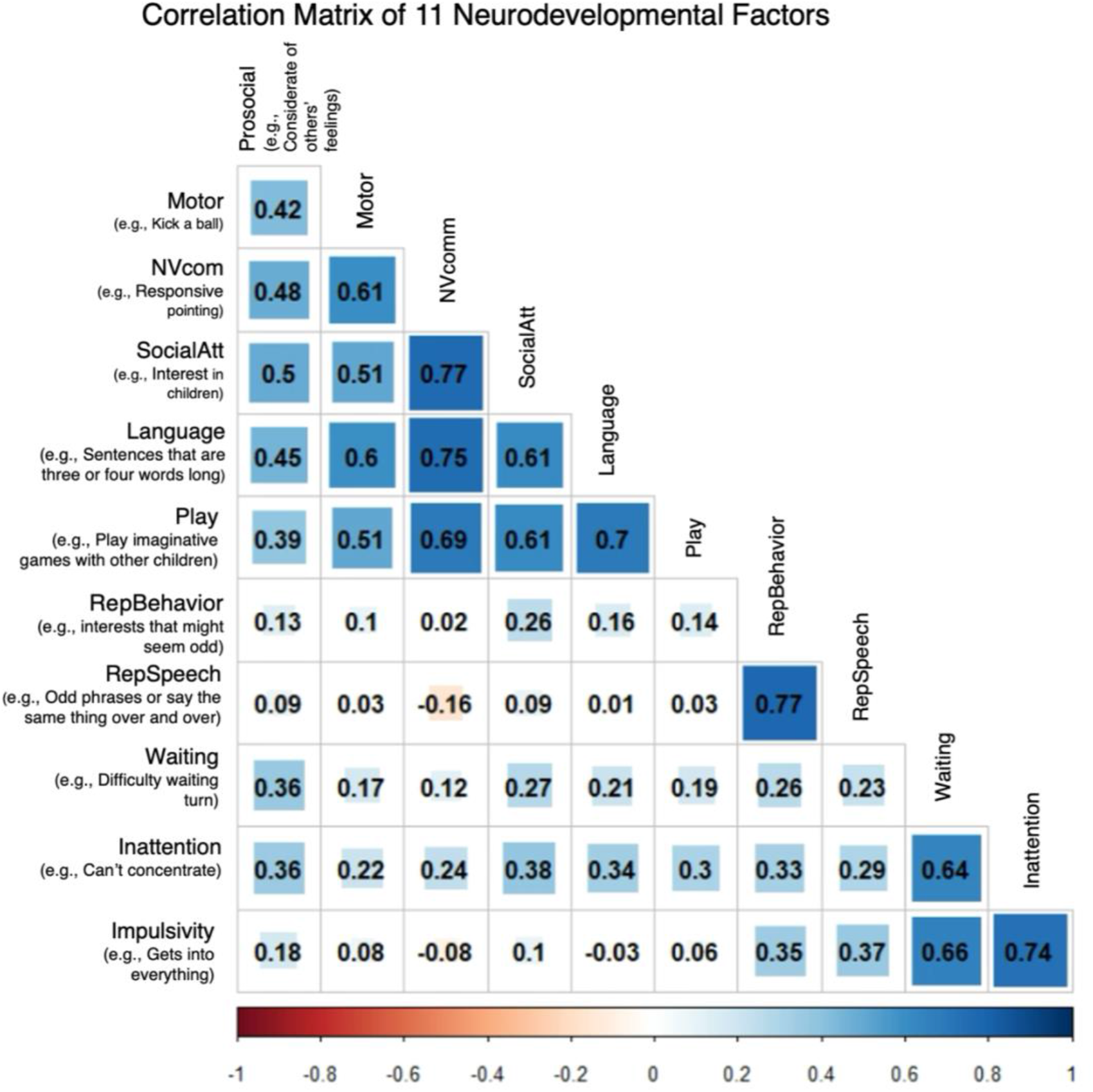
Correlation matrix of the 11 factors from the correlated factor model in the full population.

The 11-factor model included factors roughly corresponding to areas of prosocial behavior (prosocial), motor development (motor), nonverbal communication and joint attention (NVcom), social attention and interest (SocialAtt), language and verbal communication (language), play, repetitive and restricted behaviors and interests (RepBehavior), repetitive and idiosyncratic speech (RepSpeech), waiting, inattention and overactivity (inattention), and impulsivity. Most items (73/76) loaded well (λ > 0.4) onto their respective factors (Supplementary Figure S2) and all factors except the idiosyncratic speech and impulsivity factors had moderate to high positive correlations with most other factors (Figure 2). Factors covering the broad domains of social/communication, ADHD traits, and repetitive behaviors and speech were highly correlated amongst themselves but showed differing patterns of correlation with factors outside their broad domains. Parameter estimates of the final model are presented in Supplementary Tables S21-24. Finally, measurement invariance testing showed that invariance of thresholds and loadings held so factors were assumed to be largely representing the same constructs between males and females (Supplementary Table S25).

An additional general factor explaining all covariance between the different factors of early neurodevelopment had poor model fit indices (Hierarchical CFI: 0.621, TLI: 0.609, RMSEA: 0.032; Bifactor: CFI: 0.644, TLI: 0.624, RMSEA: 0.031) compared with the correlated factor model (CFI: 0.888, TLI: 0.883, RMSEA: 0.018) in the full sample. Besides fit indices, anomalous results in parameter estimates, non-uniform (λ = 0.07-0.89; Table S26) loadings, and several specific factors with variances estimated close to zero (Supplementary Tables S27) indicated misspecification of the bifactor model to the data. This was less apparent in the hierarchical model (Supplementary Tables S28-29); therefore, it was used for further analyses. However, the general factor still exhibited varied loadings (λ = 0.313 – 0.787) and was characterized by factors encompassing social, communication, and motor development, which all had strong loadings from items with low endorsement in the general population.

#### Factor validation and correlations with later outcomes

We found that nearly all early neurodevelopmental factors were associated with receiving diagnosis of any of the neurodevelopmental conditions, higher perceived impact in daily life at ages 5 and 8, later psychiatric inpatient services, and reported early referral to habilitation, special education, and psychiatric services (Supplementary Figures S3-5). In a multivariate model all outcomes were still associated with at least one factor or group of highly correlated factors, and many were associated with multiple (Figure 3; Supplementary Figures S6-7). For example, both the highly correlated groups of the ADHD-trait factors, and social and communication factors were still associated with later receiving a diagnosis of ADHD. Some of these associations also differed by sex, such as the motor factor being associated with an autism diagnosis only in girls in the multivariate model.

**Figure 3:**
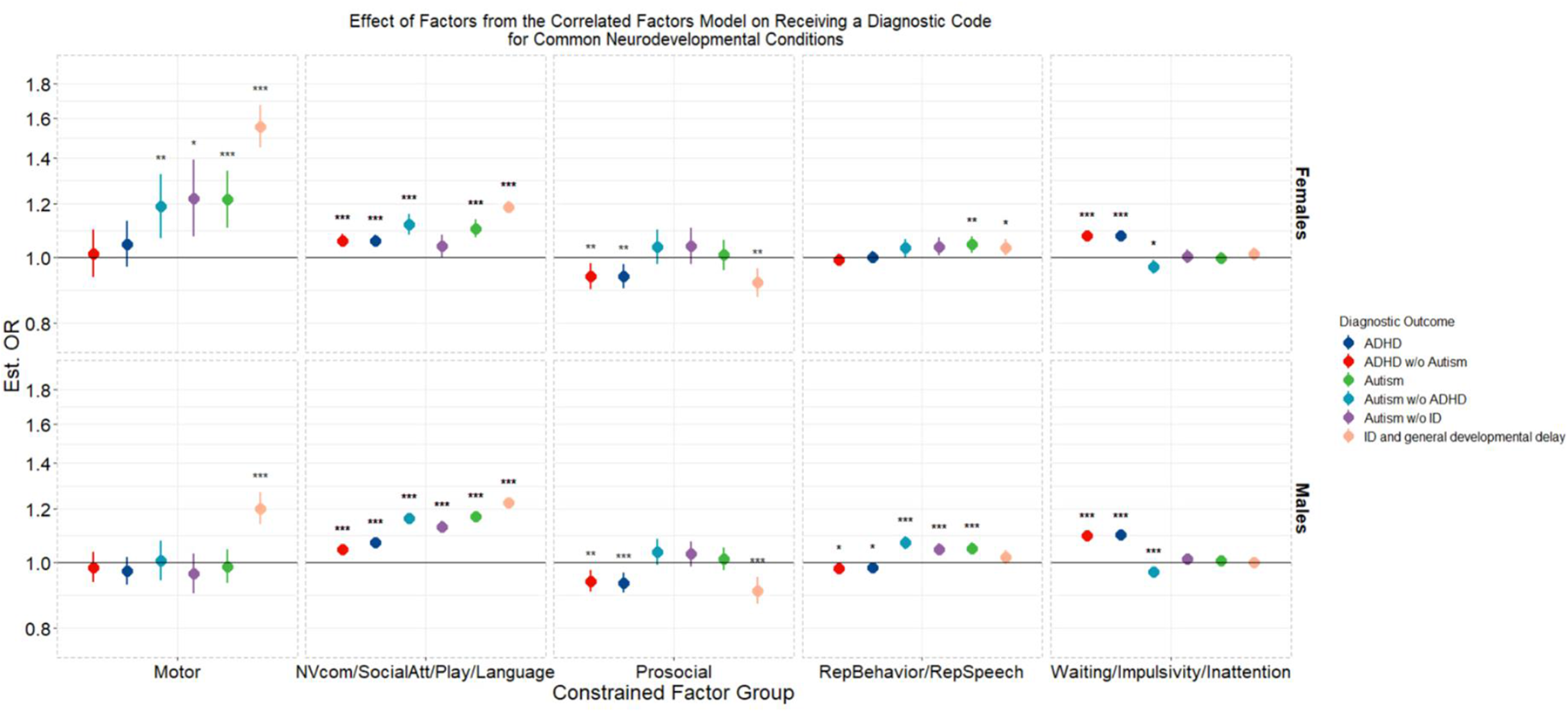
Estimated effects of factors from the correlated factor model in a multivariate regression controlling for the effects of all factors on the outcome for 5 selected diagnostic outcomes. Effects are presented as odds ratios calculated from the exponential of the standardized beta value from the logistic regression in the measurement models. 95% percent confidence intervals are shown. Due to high correlations amongst domains in the broad areas of social communication (the language & verbal communication, nonverbal communication and joint attention, play, and social attention and interest factors), ADHD-associated traits (the inattention and overactivity, waiting, impulsivity factors), and repetitive and restricted behaviors (the repetitive and idiosyncratic speech and repetitive and restricted behaviors and interests factors) effects of these factors were constrained to be equal to avoid collinearity issues.”*”, “**”, “***” denote adjusted p <0.05,<0.01,and <0.001 respectively, after multiple testing correction. For full results of the outcome models see the supplementary results.

In the hierarchical model, where specific factors simultaneously predicted the outcomes, all factors were still associated with at least one outcome and some factors within the highly correlated factor groups had differing magnitude and direction of effects from each other (Figure S8-10). For, example out of the highly correlated social and communication factors, only the play and language factors were associated with ADHD. These two factors also had the most specific factor associations, both being significantly associated with most of the outcomes. The general factor was associated with all outcomes (Supplementary Figure S8-10). However, the effect of a general factor on the outcome, when moderated by the specific factors, primarily explained additional variance in the outcomes related to early referral, general developmental delay/intellectual disability and, in girls, specific language conditions when compared to the correlated factors model (Supplementary Figure S11).

#### Common genetic variance underlying early neurodevelopmental traits

GWAS of the factor scores from the 11-factor model had low h^2^_SNP_ estimates. Four factors had estimated confidence intervals that crossed 0 (Supplementary Table S30). The highest estimate was the non-verbal communication factor (h^2^_SNP_ *=* 0.037 [0.013 – 0.061], p = 0.003). There were four unique genome-wide significant loci identified across the factors (Supplementary Tables S31-35). Three of these SNPs were associated with multiple factors (rs61775569, rs12967622, rs10956955 in LD with rs4961212, Table S36). Results from gene-based association analyses implemented in FUMA, identified three genes associated with specific factors (p < 2.682 x 10^-6^; Tables S37-47). [60] The motor factor was associated with *CNGB3* (p = 1.53 x 10^-6^), while the prosocial behavior factor was associated with *RSRC1* (p = 3.95 x 10^-7^) and *ADAMTS17* (p = 8.19 x 10^-7^). Sex-stratified factor GWAS were underpowered but showed high genetic correlation with the factors in the full sample. These GWAS showed some differences in h^2^_SNP_ estimates by sex, but these differences did not reach statistical significance (Supplementary Table S48). 34 item GWAS reached our greater than 1 h^2^_SNP_ Z threshold (Supplementary Table S49), of which, 21 items had h^2^_SNP_ that reached statistical significance. These items had a large range of estimated h^2^_SNP_ (range: 0.02 – 0.27; Supplementary Table S50) with differing levels of precision.

#### Early neurodevelopmental traits relationships with genetic liability for neurodevelopmental conditions

Genetic correlations between early neurodevelopmental traits and neurodevelopmental conditions were observed across multiple domains and were evident at both the factor and item-level (Figure 4, Supplementary Tables S51-52). ADHD had the highest genetic correlation with the inattentive and overactivity factor (*r*_g_ = 0.95 [0.13 – 1]). The prosocial behavior factor had the highest significant association for both autism (*r*_g_ = 0.56 [0.29 – 0.83]) and schizophrenia (*r*_g_ = 0.20 [0.05 – 0.34]). While common genetic variance for neurodevelopmental conditions appeared to be broadly correlated across a range of different developmental areas in early childhood there were some instances of differing effects across conditions. The strongest example of this was the positive genetic correlation between the motor factor and autism (*r*_g_ = 0.42 [0.11 – 0.72]), and to a lesser extent, schizophrenia (*r*_g_ = 0.17 [0 – 0.34]) but a negative correlation with ADHD (*r*_g_ = -0.32 [-0.58 – -0.01]).

The factors from the sex-stratified GWAS displayed similar genetic correlations with the neurodevelopmental conditions as in the entire sample but with slightly higher correlation estimates in males than females with autism and slightly higher in females than males with schizophrenia (Supplementary Table S53-54). These correlations were accompanied by large, overlapping confidence intervals but were in accordance with findings of the PGS analyses. In these analyses, effects surviving multiple testing corrections were found exclusively in males for the autism PGS and in females for the schizophrenia PGS (Supplementary Figures S12-13 and Tables S55-56).

At the item-level, the item “*considerate of feelings”* had the highest genetic correlation with autism, the item “*can’t sit still, restless or overactive”* with ADHD, and “*volunteers”* with schizophrenia (Supplementary Table S52). A few item GWAS had differing effects compared to their specified factor’s GWAS. For example, the item measuring “*excessive talking”,* which was a part of the CBCL and loaded onto the impulsivity factor, was significantly negatively correlated (*r*_g_ = -0.25 [-0.37 – -0.124]) with schizophrenia after multiple testing corrections while the impulsivity factor was uncorrelated with schizophrenia (*r*_g_ = -0.01[-0.17 – 0.15]).

**Figure 4:**
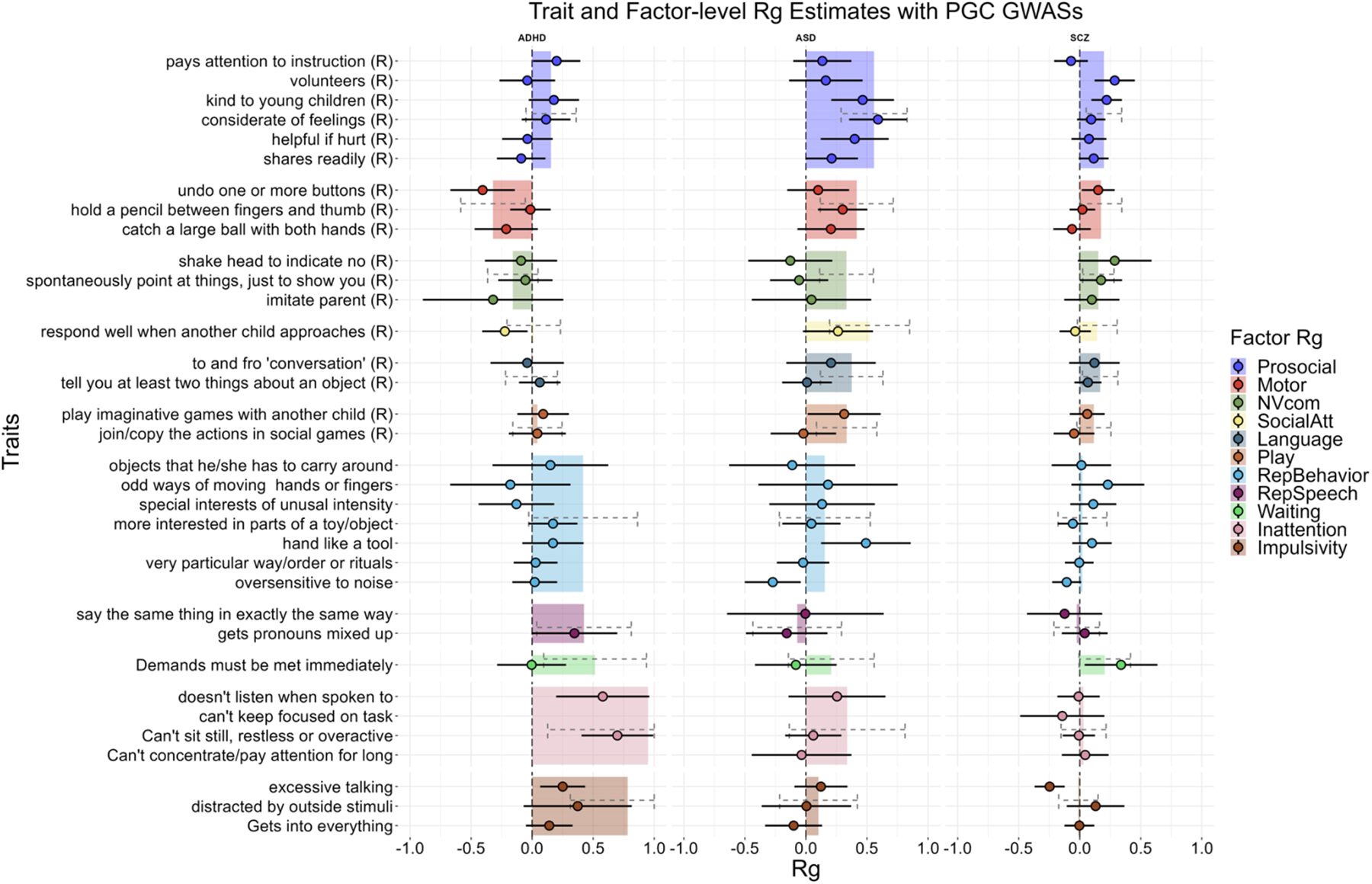
Estimated item and factor loading GWAS genetic correlation with PGS GWAS. 95% percent confidence intervals are presented. Results of multiple testing corrections are presented in Supplementary Tables S51 and S52 as a reference for the strength of statistical significance. Items are represented by points and factors bars. Bar width only reflects the number of items from that factor that were included. (R) denotes reversed coded items. The inattention factor had an estimated genetic correlation above one but is shown at 1.0. This factor as well as the impulsivity factor had upper bounds of the confidence interval estimated over 1. Item-level estimates were removed if confidence intervals were estimated as having a range larger than 1.5.

#### Genomic structure modeling and specificity of SNP effects

Given power constraints, the EFA was run on the smoothed estimated genetic correlation matrix of all chromosomes and no further downstream analyses were done. Genetic correlations between all items were estimated and are presented in Figure 5. Two to three clusters of items seem to emerge from this, the most obvious being the prosocial behavior items and the item “*uses hand like a tool.*” These items were notably the items with the highest genetic correlations with autism. The other two possible clusters were made up of items covering ADHD traits, repetitive and restricted behaviors and interests, and play behaviors and thus less interpretable. The traditional eigenvalues method indicated eight factors to be extracted in an EFA at the genomic level. Extracting one “general” factor in the EFA left many items unrepresented and was mainly defined by the “*uses hand like a tool*” and prosocial behavior items, further extraction of factors beyond this were hard to interpret and frequently had factors defined by a few items, frequent cross loading, and strong negative loadings.

The motor, prosocial behavior, RepBehavior, and inattention factors were recreated via a CFA at the genomic level. Among these, only the prosocial behavior factor demonstrated an exceptional fit (CFI = 1, SRMR = 0.095; Supplementary Figure S14) and exhibited strong and significant loadings for most items. Only one item, specifically “*pays attention to your instructions*” did not exhibit a significant loading on this factor. This item differed from the rest as it was not part of the SDQ scale but was asked alongside the SDQ in the MoBa questionnaire. The inattention and overactivity factor had significant loadings for all items but as it was comprised of three items, fit indices could not be estimated. The repetitive and restricted behaviors and interests had excellent fit (CFI = 0.99, SRMR = 0.083) but non-significant loadings for all items. The motor factor did not converge.

Based on the outcomes of the common factor models, we only performed a subsequent common factor GWAS for the prosocial behavior factor. This did not yield any genome-wide significant loci but identified 6 independent SNPs hits and 17 Q_snp_ at a suggestive association threshold (p < 5 x 10^-5^; Supplementary Tables S57-58). Removing the item that did not significantly load onto the common genetic factor, the common factor GWAS identified 7 SNPs and 9 Q_snp_ at the same threshold (Supplementary Figure S15; Supplementary Tables S59-60).

**Figure 5:**
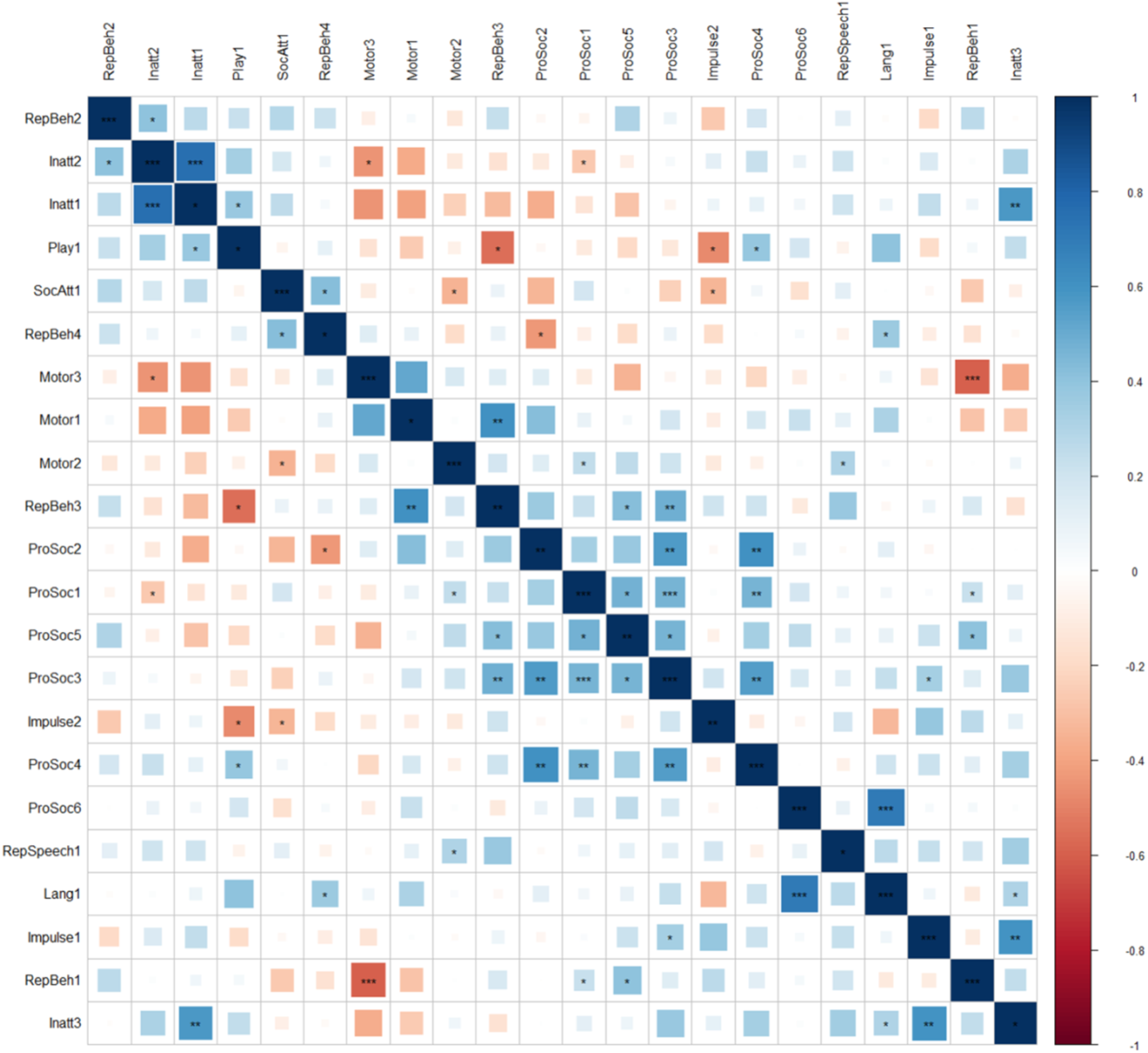
The estimated smoothed genetic correlations matrix for the 22 neurodevelopmental items used in the EFA and genetic factor modeling. Items order using angular order of the eigenvectors (AOE).” *”, “**”, “***” denote uncorrected p <0.05, <0.01, and <0.001 respectively.

## Discussion

We leveraged the item-level questionnaire data in n= 58 630 MoBa children to investigate patterns of relationships between specific traits from different areas of development in early childhood, the underlying genetic contributions, and potential shared etiology to clinically diagnosed neurodevelopmental conditions. The main findings are firstly, a high heterogeneity at both the phenotypic and genotypic level underlying early neurodevelopmental traits – higher than would be expected if these traits were neatly aligned with distinct neurodevelopmental conditions. Secondly, despite their etiological and structural heterogeneity, early neurodevelopmental traits in a general population sample *are* phenotypically and genetically associated with neurodevelopmental diagnoses.

### Heterogeneity underlying early neurodevelopmental traits in a population-based sample

We find that most domains of neurodevelopment traits are at least moderately correlated with each other at the phenotypic level. The simplest etiological explanation for this would be shared liability across all areas of neurodevelopment, such as a general genetic neurodevelopment factor, which has been suggested based on twin studies and clinical observations [17,63,64]. However, we also show substantial heterogeneity underlying early neurodevelopmental traits both at the phenotypic and genetic levels, and little evidence supporting a general factor of liability to all early neurodevelopmental traits at either level of analysis. Besides poor model fit of the hierarchical and bifactor models, the general factor was not so general, indexed primarily by the specific factors or items in the factors of nonverbal communication and social attention. On the genotypic level, we observed considerable range in the magnitude and direction of genetic correlations between items. Further, the EFA at the genotypic level did not provide support for a single factor of common genetic liability.

Notwithstanding the question of the existence of a general factor for neurodevelopmental traits, we observe increased heterogeneity compared to what would be expected based on etiological factors that neatly lined up with diagnostic criteria. The observed factors are highly correlated amongst themselves in domains related to commonly separated neurodevelopmental domains (i.e., social and communication, repetitive behaviors, ADHD traits). However, these factors are differentially correlated with the factors outside of their domain and, when correlations between factors were accounted for by a general factor, have differing associations with later diagnoses. To note, while the neurodevelopmental traits are associated with and share some genetic variance with neurodevelopmental conditions, traits are non-specific to conditions. This pattern is consistent with co-occurrence between neurodevelopmental conditions being commonplace – in many cases being the rule, rather than the exception [13,64,65].

While limited by power, the results of the genomic factor modeling points towards a similar level of heterogeneity in the genetic architecture of early neurodevelopmental traits. We find some evidence for common genetic factors that resemble the prosocial and the inattention factors identified in the phenotypic models. This is supported by the good fit indices and/or significant loadings in the genomic common factor models. Shared genome-wide significant SNPs across the factor GWAS indicates the existence of some shared genetic loci, particularly across social and communication factors and prosocial factor. However, in other areas such as motor development, increased heterogeneity is observed. While the motor factor shares a genome-wide significant locus with nonverbal communication and joint attention, the factor is defined by items covering gross motor skills. At the item-level, one fine motor item does not exhibit strong correlation with the other two motor items and the motor genomic common factor model does not converge. This potentially suggesting the presence of different genetic mechanisms underlying different aspects of motor skills. Lastly, even among the items measuring prosocial behavior, the higher number of Q_SNP_ hits compared to SNP hits contributing to the common genetic factor at a suggestive association threshold emphasizes the possibility of item-level specificity of genetic effects, even within the most coherent genetic factor.

### Early neurodevelopmental traits are associated with neurodevelopmental conditions

The factors identified as underpinning early neurodevelopmental traits in our sample were associated with receiving a clinical diagnosis for different neurodevelopmental conditions. We generally find stronger associations between conditions and factors that contain items that overlap with diagnostic criteria of that condition such as the social and communication factors with autism or the ADHD trait factors with ADHD. Stronger associations are also seen for conditions that have higher rates of earlier referral or diagnosis in our sample such as intellectual disability and specific motor conditions. The strength of these associations is likely impacted by the overlap in items with diagnostic criteria, however we still find associations of factors with conditions with later age of onset, such as specific learning conditions as well as with conditions which do not have diagnostic criteria overlapping with the factor, such as the social and communication factors with ADHD even after excluding individuals who have also received an autism diagnosis.

Early neurodevelopmental traits shared common genetic liability with ADHD, autism, and schizophrenia. While limited by power, many items suggest similar associations to neurodevelopmental conditions as their factor. However, we do observe some trait-level heterogeneity as in the recent item-level genomic analysis of neuroticism [66]. For example, while the prosocial factor was genetically correlated with autism and schizophrenia genetic liability, only the factor’s items “*kind to young children*” and “*considerate of feelings”* had associations with autism and only the items “*volunteers”* and “*kind to young children*” with schizophrenia after multiple testing corrections. Childhood prosocial behavior has been previously associated with schizophrenia polygenic liability; however, this same study did not find an association with autism genetic risk [67]. Additionally showing trait-level heterogeneity, items in the motor and repetitive behavior factors have a range in the magnitude and sometimes direction of genetic correlations with neurodevelopmental conditions. These observations offer some potential areas for follow up work in clinical samples identifying differentiating mechanisms of early development across conditions.

Our findings also identify some potential for shared mechanisms across domains at the sub-diagnostic level. For instance, the repetitive behavior and repetitive speech factors are more correlated with the ADHD trait factors than some of the social and communication factors and had higher genetic correlation estimates with ADHD than with autism. While this observation may offer areas of potential shared early signs across conditions, the validity of items in a general population should also be considered. For example, items such as “*says the same thing over and over*” could be misinterpreted by parents, resulting in it capturing activity level or more common behaviors, rather than the idiosyncratic speech typically associated with autism. This may also contribute to the low (or lack of) estimated heritability in some of these items.

### Limitations

There are some limitations of our study that should be considered. Despite splitting our sample into discovery and test halves, the exploratory factor analysis of such a diverse set of items, in a large sample, is likely to have led to some level of overfitting. Because of this, we do not suggest interpreting all identified factors as necessarily definitive distinct factors with important etiological meaning but instead put forward that there is increased dimensionality across areas of development with differing relationships to each other and to neurodevelopmental conditions that may be lost at diagnostic or scale level.

Low power to detect signal for many of the item GWAS limits the claims. For the effects we identified the increased variation due to underpowered GWAS may contribute to the large range in estimated genetic correlations. However, power concerns are unlikely to fully explain the lack of a single general genetic factor. Our genetic analyses were also limited to common genetic variants.

There is considerable overlap of rare variants associated with different neurodevelopmental conditions [22–24] and rare variants are more common in cases presenting with intellectual disability and/or developmental delays [68,69], which the general factor in the phenotypic model was most associated with in the diagnostic outcome analyses. Finally, the genetic analyses were limited to participants in MoBa of European genetic ancestry, limiting the generalizability of our results across ancestries.

### Conclusions

Our exploratory results reveal the multidimensionality underlying early neurodevelopmental traits in a population-based birth cohort. These dimensions are broadly associated with receiving a diagnosis of neurodevelopmental conditions, and many genetically correlated with ADHD, autism, and/or schizophrenia. We find little support for a shared common genetic liability across all traits in the general population. Instead, we observe multiple specific factors with certain shared genetic loci identified across, particularly, the social and communication domains of neurodevelopment, but none that are evidently relevant across all domains. Our trait-level analyses highlight the role of heterogenous genetic effects underlying early neurodevelopment traits and their relationships to neurodevelopmental conditions. These findings provide areas for further investigation to identify shared and distinct mechanisms across neurodevelopmental conditions.

## Data Availability

The consent given by the participants does not allow for storage of data on an individual level in repositories. Researchers can apply for access to data for replication purposes via MoBa, in line with MoBa data access policies.

## Acknowledgements

The Norwegian Mother, Father and Child Cohort Study is supported by the Norwegian Ministry of Health and Care Services and the Ministry of Education and Research. We are grateful to all the participating families in Norway who take part in this on-going cohort study. We thank the Norwegian Institute of Public Health (NIPH) for generating high-quality genomic data. This research is part of the HARVEST collaboration, supported by the Research Council of Norway (#229624). We also thank the NORMENT Centre for providing genotype data, funded by the Research Council of Norway (#223273), South East Norway Health Authorities and Stiftelsen Kristian Gerhard Jebsen. We further thank the Center for Diabetes Research, the University of Bergen for providing genotype data and performing quality control and imputation of the data funded by the ERC AdG project SELECTionPREDISPOSED, Stiftelsen Kristian Gerhard Jebsen, Trond Mohn Foundation, the Research Council of Norway, the Novo Nordisk Foundation, the University of Bergen, and the Western Norway Health Authorities.

This work was performed on the TSD (Tjeneste for Sensitive Data) facilities, owned by the University of Oslo, operated and developed by the TSD service group at the University of Oslo, IT Department (USIT). Analyses were performed on resources provided by Sigma2 – the National Infrastructure for High-Performance Computing and Data Storage in Norway.

Disclaimer. Data from the Norwegian Patient Registry has been used in this publication. The interpretation and reporting of these data are the sole responsibility of the authors, and no endorsement by the Norwegian Patient Registry is intended nor should be inferred.

## References

1. Diagnostic and statistical manual of mental disorders. 5th ed. American Psychiatric Association; 2013.

2. ICD-11. Int Classif Dis 11th Revis. 2018;

3. Nikolas M, Burt A. Genetic and environmental influences on ADHD symptom dimensions of inattention and hyperactivity: A meta-analysis. – PsycNET. J Abnorm Psychol. 2010;119(1):1–17.

4. Lichtenstein P, Carlström E, Råstam M, Gillberg C, Anckarsäter H. The Genetics of Autism Spectrum Disorders and Related Neuropsychiatric Disorders in Childhood. Am J Psychiatry. 2010 Nov 1;167(11):1357–63.

5. Tick B, Bolton P, Happé F, Rutter M, Rijsdijk F. Heritability of autism spectrum disorders: a meta-analysis of twin studies. J Child Psychol Psychiatry. 2016;57(5):585–95.

6. Nigg JT, Sibley MH, Thapar A, Karalunas SL. Development of ADHD: Etiology, Heterogeneity, and Early Life Course. Annu Rev Dev Psychol. 2020;2(1):559–83.

7. Lord C, Brugha TS, Charman T, Cusack J, Dumas G, Frazier T, et al. Autism spectrum disorder. Nat Rev Dis Primer. 2020 Jan 16;6(1):1–23.

8. Loomes R, Hull L, Mandy WPL. What Is the Male-to-Female Ratio in Autism Spectrum Disorder? A Systematic Review and Meta-Analysis. J Am Acad Child Adolesc Psychiatry. 2017 Jun 1;56(6):466–74.

9. Willcutt EG. The Prevalence of DSM-IV Attention-Deficit/Hyperactivity Disorder: A Meta-Analytic Review. Neurotherapeutics. 2012 Jul 1;9(3):490–9.

10. Quinn JM, Wagner RK. Gender Differences in Reading Impairment and in the Identification of Impaired Readers: Results From a Large-Scale Study of At-Risk Readers. J Learn Disabil. 2015 Jul 1;48(4):433–45.

11. Thapar A, Cooper M, Rutter M. Neurodevelopmental disorders. Lancet Psychiatry. 2017 Apr;4(4):339–46.

12. Lundström S, Reichenberg A, Melke J, Råstam M, Kerekes N, Lichtenstein P, et al. Autism spectrum disorders and coexisting disorders in a nationwide Swedish twin study. J Child Psychol Psychiatry. 2015;56(6):702–10.

13. Brimo K, Dinkler L, Gillberg C, Lichtenstein P, Lundström S, Åsberg Johnels J. The co-occurrence of neurodevelopmental problems in dyslexia. Dyslexia. 2021;27(3):277–93.

14. Reiersen AM, Constantino JN, Volk HE, Todd RD. Autistic traits in a population-based ADHD twin sample. J Child Psychol Psychiatry. 2007;48(5):464–72.

15. Jokiranta-Olkoniemi E, Cheslack-Postava K, Joelsson P, Suominen A, Brown AS, Sourander A. Attention-deficit/hyperactivity disorder and risk for psychiatric and neurodevelopmental disorders in siblings. Psychol Med. 2019 Jan;49(1):84–91.

16. Jokiranta-Olkoniemi E, Cheslack-Postava K, Sucksdorff D, Suominen A, Gyllenberg D, Chudal R, et al. Risk of Psychiatric and Neurodevelopmental Disorders Among Siblings of Probands With Autism Spectrum Disorders. JAMA Psychiatry. 2016 Jun 1;73(6):622–9.

17. Pettersson E, Anckarsäter H, Gillberg C, Lichtenstein P. Different neurodevelopmental symptoms have a common genetic etiology. J Child Psychol Psychiatry. 2013;54(12):1356–65.

18. Demontis D, Walters RK, Martin J, Mattheisen M, Als TD, Agerbo E, et al. Discovery of the first genome-wide significant risk loci for attention deficit/hyperactivity disorder. Nat Genet. 2019 Jan;51(1):63–75.

19. Grove J, Ripke S, Als TD, Mattheisen M, Walters RK, Won H, et al. Identification of common genetic risk variants for autism spectrum disorder. Nat Genet. 2019 Mar;51(3):431–44.

20. Doust C, Fontanillas P, Eising E, Gordon SD, Wang Z, Alagöz G, et al. Discovery of 42 genome-wide significant loci associated with dyslexia. Nat Genet. 2022 Nov;54(11):1621–9.

21. Grotzinger AD, Mallard TT, Akingbuwa WA, Ip HF, Adams MJ, Lewis CM, et al. Genetic architecture of 11 major psychiatric disorders at biobehavioral, functional genomic and molecular genetic levels of analysis. Nat Genet. 2022 May;54(5):548–59.

22. Satterstrom FK, Walters RK, Singh T, Wigdor EM, Lescai F, Demontis D, et al. Autism spectrum disorder and attention deficit hyperactivity disorder have a similar burden of rare protein-truncating variants. Nat Neurosci. 2019 Dec;22(12):1961–5.

23. Satterstrom FK, Kosmicki JA, Wang J, Breen MS, Rubeis SD, An JY, et al. Large-Scale Exome Sequencing Study Implicates Both Developmental and Functional Changes in the Neurobiology of Autism. Cell. 2020 Feb 6;180(3):568–584.e23.

24. Martin J, Cooper M, Hamshere ML, Pocklington A, Scherer SW, Kent L, et al. Biological Overlap of Attention-Deficit/Hyperactivity Disorder and Autism Spectrum Disorder: Evidence From Copy Number Variants. J Am Acad Child Adolesc Psychiatry. 2014 Jul 1;53(7):761–770.e26.

25. Ismail FY, Shapiro BK. What are neurodevelopmental disorders? Curr Opin Neurol. 2019 Aug;32(4):611–6.

26. Reiersen A. How should we classify complex neurodevelopmental disorders? Scand J Child Adolesc Psychiatry Psychol. 2017 Jul 5;5.

27. Mullin AP, Gokhale A, Moreno-De-Luca A, Sanyal S, Waddington JL, Faundez V. Neurodevelopmental disorders: mechanisms and boundary definitions from genomes, interactomes and proteomes. Transl Psychiatry. 2013 Dec;3(12):e329–e329.

28. Taurines R, Schwenck C, Westerwald E, Sachse M, Siniatchkin M, Freitag C. ADHD and autism: differential diagnosis or overlapping traits? A selective review. ADHD Atten Deficit Hyperact Disord. 2012 Sep 1;4(3):115–39.

29. Robinson EB, St Pourcain B, Anttila V, Kosmicki JA, Bulik-Sullivan B, Grove J, et al. Genetic risk for autism spectrum disorders and neuropsychiatric variation in the general population. Nat Genet. 2016 May;48(5):552–5.

30. Hannigan LJ, Askeland RB, Ask H, Tesli M, Corfield E, Ayorech Z, et al. Genetic Liability for Schizophrenia and Childhood Psychopathology in the General Population. Schizophr Bull. 2021 Feb 9;47(4):1179–89.

31. Martin J, Hamshere ML, Stergiakouli E, O’Donovan MC, Thapar A. Genetic risk for attention-deficit/hyperactivity disorder contributes to neurodevelopmental traits in the general population. Biol Psychiatry. 2014 Oct 15;76(8):664–71.

32. Stergiakouli E, Martin J, Hamshere ML, Langley K, Evans DM, St Pourcain B, et al. Shared genetic influences between attention-deficit/hyperactivity disorder (ADHD) traits in children and clinical ADHD. J Am Acad Child Adolesc Psychiatry. 2015 Apr;54(4):322–7.

33. Askeland RB, Hannigan LJ, Ask H, Ayorech Z, Tesli M, Corfield E, et al. Early manifestations of genetic risk for neurodevelopmental disorders. J Child Psychol Psychiatry. 2022 Jul;63(7):810–9.

34. Thomas TR, Koomar T, Casten LG, Tener AJ, Bahl E, Michaelson JJ. Clinical autism subscales have common genetic liabilities that are heritable, pleiotropic, and generalizable to the general population. Transl Psychiatry. 2022 Jun 13;12(1):1–14.

35. van ’t Hof M, Tisseur C, van Berckelear-Onnes I, van Nieuwenhuyzen A, Daniels AM, Deen M, et al. Age at autism spectrum disorder diagnosis: A systematic review and meta-analysis from 2012 to 2019. Autism. 2021 May 1;25(4):862–73.

36. Knight T, Steeves T, Day L, Lowerison M, Jette N, Pringsheim T. Prevalence of Tic Disorders: A Systematic Review and Meta-Analysis. Pediatr Neurol. 2012 Aug 1;47(2):77–90.

37. Rocco I, Corso B, Bonati M, Minicuci N. Time of onset and/or diagnosis of ADHD in European children: a systematic review. BMC Psychiatry. 2021 Nov 16;21(1):575.

38. Magnus P, Irgens LM, Haug K, Nystad W, Skjærven R, Stoltenberg C, et al. Cohort profile: The Norwegian Mother and Child Cohort Study (MoBa). Int J Epidemiol. 2006 Oct 1;35(5):1146–50.

39. Magnus P, Birke C, Vejrup K, Haugan A, Alsaker E, Daltveit AK, et al. Cohort Profile Update: The Norwegian Mother and Child Cohort Study (MoBa). Int J Epidemiol. 2016 Apr 1;45(2):382–8.

40. 40. Corfield EC, Frei O, Shadrin AA, Rahman Z, Lin A, Athanasiu L, et al. The Norwegian Mother, Father, and Child cohort study (MoBa) genotyping data resource: MoBaPsychGen pipeline v.1 [Internet]. bioRxiv; 2022 [cited 2023 Feb 2]. p. 2022.06.23.496289. Available from: https://www.biorxiv.org/content/10.1101/2022.06.23.496289v3

41. Rutter M, Lord C, Bailey A. SCQ The Social Communication Questionnaire: Manual. Los Angel West Psychol Serv. 2003b;

42. Squires J, Potter L, Brikker D. The ASQ User’s Guide. 2nd ed. Baltimore: Paul H. Brookes Publishing Co; 1999.

43. Schjolberg S. Early Identification of Autism Spectrum Disorders. The Social Brain; 2003; Gøteborg.

44. Robins DL, Fein D, Barton ML, Green JA. The Modified Checklist for Autism in Toddlers: an initial study investigating the early detection of autism and pervasive developmental disorders. J Autism Dev Disord. 2001 Apr;31(2):131–44.

45. Swinkels SHN, Dietz C, van Daalen E, Kerkhof IHGM, van Engeland H, Buitelaar JK. Screening for autistic spectrum in children aged 14 to 15 months. I: the development of the Early Screening of Autistic Traits Questionnaire (ESAT). J Autism Dev Disord. 2006 Aug;36(6):723–32.

46. Achenbach TM. Manual for child behavior checklist. Scientific Research Publishing; 1992.

47. Goodman R. The Strengths and Difficulties Questionnaire: A Research Note. J Child Psychol Psychiatry. 1997;38(5):581–6.

48. Hannigan L. MoBa Phenotools [Internet]. 2023. Available from: https://github.com/psychgen/phenotools

49. Choi SW, O’Reilly PF. PRSice-2: Polygenic Risk Score software for biobank-scale data. GigaScience. 2019 Jul 1;8(7):giz082.

50. Trubetskoy V, Pardiñas AF, Qi T, Panagiotaropoulou G, Awasthi S, Bigdeli TB, et al. Mapping genomic loci implicates genes and synaptic biology in schizophrenia. Nature. 2022 Apr;604(7906):502–8.

51. Murray RM, Bhavsar V, Tripoli G, Howes O. 30 Years on: How the Neurodevelopmental Hypothesis of Schizophrenia Morphed Into the Developmental Risk Factor Model of Psychosis. Schizophr Bull. 2017 Oct 21;43(6):1190–6.

52. Insel TR. Rethinking schizophrenia. Nature. 2010 Nov 11;468(7321):187–93.

53. Morris-Rosendahl DJ, Crocq MA. Neurodevelopmental disorders—the history and future of a diagnostic concept. Dialogues Clin Neurosci. 2020 Mar 31;22(1):65–72.

54. Coombes BJ, Ploner A, Bergen SE, Biernacka JM. A principal component approach to improve association testing with polygenic risk scores. Genet Epidemiol. 2020;44(7):676–86.

55. Wu H, Estabrook R. Identification of Confirmatory Factor Analysis Models of Different Levels of Invariance for Ordered Categorical Outcomes. Psychometrika. 2016 Dec;81(4):1014–45.

56. Rosseel Y. lavaan: An R Package for Structural Equation Modeling. J Stat Softw [Internet]. 2012 [cited 2023 Jan 30];48(2). Available from: http://www.jstatsoft.org/v48/i02/

57. Jorgensen TD, Pornprasertmanit S, Schoemann AM, Rosseel Y. semTools: Useful tools for structural equation modeling [Internet]. 2022. Available from: https://CRAN.R-project.org/package=semTools

58. Mbatchou J, Barnard L, Backman J, Marcketta A, Kosmicki JA, Ziyatdinov A, et al. Computationally efficient whole-genome regression for quantitative and binary traits. Nat Genet. 2021 Jul;53(7):1097–103.

59. Bulik-Sullivan B, Finucane HK, Anttila V, Gusev A, Day FR, Loh PR, et al. An atlas of genetic correlations across human diseases and traits. Nat Genet. 2015 Nov;47(11):1236–41.

60. Watanabe K, Taskesen E, van Bochoven A, Posthuma D. Functional mapping and annotation of genetic associations with FUMA. Nat Commun. 2017 Nov 28;8(1):1826.

61. R Core Team. R: A Language and Environment for Statistical Computing [Internet]. Vienna, Austria: R Foundation for Statistical Computing; 2021. Available from: https://www.R-project.org/

62. Grotzinger AD, Rhemtulla M, de Vlaming R, Ritchie SJ, Mallard TT, Hill WD, et al. Genomic structural equation modelling provides insights into the multivariate genetic architecture of complex traits. Nat Hum Behav. 2019 May;3(5):513–25.

63. Moreno-De-Luca A, Myers SM, Challman TD, Moreno-De-Luca D, Evans DW, Ledbetter DH. Developmental brain dysfunction: revival and expansion of old concepts based on new genetic evidence. Lancet Neurol. 2013 Apr;12(4):406–14.

64. Gillberg C. The ESSENCE in child psychiatry: Early Symptomatic Syndromes Eliciting Neurodevelopmental Clinical Examinations. Res Dev Disabil. 2010 Nov;31(6):1543–51.

65. Kaplan BJ, Dewey DM, Crawford SG, Wilson BN. The term comorbidity is of questionable value in reference to developmental disorders: data and theory. J Learn Disabil. 2001;34(6):555–65.

66. Nagel M, Watanabe K, Stringer S, Posthuma D, van der Sluis S. Item-level analyses reveal genetic heterogeneity in neuroticism. Nat Commun. 2018 Mar 2;9(1):905.

67. Schlag F, Allegrini AG, Buitelaar J, Verhoef E, van Donkelaar M, Plomin R, et al. Polygenic risk for mental disorder reveals distinct association profiles across social behaviour in the general population. Mol Psychiatry. 2022 Mar;27(3):1588–98.

68. Girirajan S, Brkanac Z, Coe BP, Baker C, Vives L, Vu TH, et al. Relative burden of large CNVs on a range of neurodevelopmental phenotypes. PLoS Genet. 2011 Nov;7(11):e1002334.

69. Warrier V, Zhang X, Reed P, Havdahl A, Moore TM, Cliquet F, et al. Genetic correlates of phenotypic heterogeneity in autism. Nat Genet. 2022 Jun 2;1–12.

