## Supplemental material for "Genetic and phenotypic heterogeneity in early neurodevelopmental traits in the Norwegian Mother, Father and Child Cohort Study"

### Supplementary Methods

### Measures

*Measures used in factor models.*

Items were included that covered current development in motor, language, social, communication, attention, activity regulation, and flexibility of behaviors and interests. 5 items were selected that covered motor development, 4 from the Ages and Stages Questionnaire (ASQ) motor subscale and 1 from the Child Behavior Checklist (CBCL). 6 items were selected that covered prosocial behavior, 5 from the Strength and Difficulties Questionnaire (SDQ) and 1 MoBa specific question. The ASQ language subscale contained 6 items measuring receptive and expressive language development. Language development as well as nonverbal communication, social behaviors, and verbal communication was also measured by the 29 social and communication question of the Social Communication Questionnaire (SCQ), 4 items from the Non-Verbal Communication Checklist (NVCC), 6 items from Modified Checklist for Autism in Toddlers (M-CHAT), 2 from Early Screening for Autistic Traits Questionnaire (ESAT), and one MoBa specific question. Flexibility of behaviors and interests was measured by the 11 repetitive and restricted behaviors and interest items from the SCQ. Attention and activity regulation were measured by 6 CBCL items and 5 MoBa specific child behavior and manners (CBM) items. Sensory perception differences were not well captured by items included in the MoBa age 3 questionnaire but 2 items, M-CAHT item "*oversensitive to noise"* and a MoBa specific item "*Has a high pain threshold"*, were included in the EFA and measured aspects of sensory differences. A full table of items, the scales they belong to, endorsement rates, and missingness is presented in Supplementary Table S1.

*Diagnostic and clinically relevant outcomes*

For diagnostic outcomes using data ascertained from the Norwegian Patient Register (NPR) individuals were included in a diagnostic outcome if they had received a diagnostic code at least one time. Further diagnostic outcome models were also run for more specific outcomes including ADHD without co-occurring autism, autism without co-occurring ADHD, autism without intellectual disability, and autism with functional speech at age 3. This last outcome was defined as receiving an autism diagnostic code and having mothers report the child having the ability to form sentences three to four words long in the age 3 questionnaire (SCQ item 1).

Clinically relevant outcomes for multimorbidity and psychiatric hospitalization were also created using data from NPR. The dichotomous variable for multimorbidity was coded so individuals were considered to have multiple neurodevelopmental diagnoses if they received at least two diagnostic codes in more than one of the main diagnostic categories. For example, receiving codes for a specific learning condition and autism would be considered multimorbidity but receiving codes for two different specific learning conditions would not. The dichotomous variable for "ever hospitalized in association with any psychiatric condition" was coded so that an individual was coded as ever being hospitalized if they had any F chapter diagnosis applicable to childhood, adolescence, and early adulthood registered in NPR during a hospitalization.

Several clinically relevant outcomes were also coded using the MoBa questionnaire data. These included measures of early service use and later maternally perceived impact and impairment from difficulties in development and behavior in their child's life. The measure of early service use was coded so that if mothers reported their child being referred to any of the following services (Habilitation service, educational psychology service, or child psychiatric clinic/department) since the age of 18$^{th}$ months, the variable was considered endorsed. The measures of impact from difficulties in development and behavior were reported in the age 5 & 8 questionnaire were collected for mothers who reported children having problems in one or more developmental or emotional/behavioral area. These items were trichotomous (no/yes, a little/ yes, a lot) and were from or based off the impact and impairment of the Strengths and Difficulties Questionnaires (SDQ). They were summed to create a total score reflecting impact across home/with family, relationships with friends, and at school. Due to the gating of the question only a small subset of the sample who had reported their child having difficulties in one or more developmental or emotional/behavioral area had scores. To distinguish between those who did not complete the 5- or 8-year questionnaires as a whole and those who did not report any difficulties the variables were recoded so that those who had questionnaire data at that timepoint, but no reported difficulties were coded as 0. Scores were collapsed into values 0-4 due to low endorsement of the highest values.

*Item GWAS*

To maximize statistical power for the item-level genome-wide association studies, trichotomous items were dichotomized so that the lowest response category was used as the reference category, collapsing the two highest response categories. For items where the lowest response category was not the most frequently endorsed, the lowest and middle response categories were collapsed and used as the reference category.

### Analyses

*Exploratory and Confirmatory Factor Analysis*

The exploratory factor analyses used in the included all items with an inter-item correlation greater than 0, this excluded two items: SCQ items "*complicated movements of their whole body*" and "*socially inappropriate questions or statements*". SCQ item "*shakes head to indicate yes*" was also excluded as it had an estimated tetrachoric correlation at 0.9 with SCQ item "*shakes head to indicate no*".

To determine the number of factors to retain we used results from examining the scree plot, parallel analysis, eigenvalues, optimal coordinates and compared alternate fit indices (RMSEA, CFI, TLI, and SRMR) after extracting 1-15 factors in the EFA based on interpretation of the scree plot. Good fit criteria were defined as CFI & TLI >= 0.95, RMSEA < 0.05, SRMR < 0.08 [1]. Interpretability of the factors, recommendation of greater than 3 items per factor [2], and parsimony of the model were also considered. EFA models were clustered on maternal ID to account for relatedness in the sample.

Models for CFA were derived from EFA models meeting the above criteria. Which factor items were determined to load onto was decided using the following criteria: First, the 3 items with the highest loading onto a factor were specified to load onto that factor; Second, remaining items were specified to load on the factor they had the highest loading onto; Third, items with loadings less than 0.3 which were not a part of the strongest three items on a factor were removed. Sensitivity analyses with removing one sibling from sibling pairs were run for the CFA analyses. Additional, sensitivity analyses were also run for several theoretically driven models allowing factors or items to correlate outside of a general factor (in g-factor models) or common factors (in correlated factor models) to assess if this explained low fit indices. Delta parameterization was used in all EFA and CFA models. Polychoric correlations were used in all factor analyses. Missing data was address using pairwise deletion for all EFA and CFA models.

Measurement invariance testing was conducted to investigate if items were capturing the same underlying constructs given the notable sex differences observed in neurodevelopmental conditions. These models were specified based on recommendations of Wu & Estabrook [3] and guidelines of Svetina, Rutkowski, and Rutkowski [4] . Following these we first specified a baseline model using equation 7 in Wu & Estabrook. This identified the model using delta parameterization with latent factor variance constrained to 1, latent factor means to 0, latent item response factor means to 0, and scale to 1 in both groups. Thresholds and loadings were freely estimated parameters in the model. To assess equality of the thresholds and loadings, we specified a model in accordance to proposition 11 in Wu & Estabrook which was equality of the thresholds, loadings, and latent item response factor means. This model was identified with latent item response factor means constrained to 0 in both groups and the latent factor variance constrained to 1, latent factor means constrained to

0, and scale constrained to 1 in the first group. Conservative criteria for change in Comparative Fit Index (ΔCFI > -Δ0.002) and McDonald´s Noncentrality Index (ΔMcNCI > - Δ0.008) between the model was used [5].

*Genetic Analyses*

GWAS of the factors and items were conducted on subsamples (n = 41708 - 42934) who had both the phenotypic data needed for the GWAS and quality controlled genotyped data available. Individual items were selected to run GWAS based on a lenient power threshold of 0.4 at an OR of 1.2, MAF of 0.01, and alpha of 0.01 in a logistic model with additive genetic effects. Power calculations were done using these parameters as well as estimating power for effects with OR at 1.1,1.15, 1, and 1.25. Power calculations were done using the R package *genpwr*. [6] Scripts for the power analyses can be found at https://github.com/psychgen/neurodevelopment_traits_structure

GWAS were run using the *Regenie* software. Factor GWAS were run using a linear regression model for association testing and for the individual item GWAS a logistic regression was used. For the item GWAS a firth correction was applied to help control biases arising from imbalanced case-control ratios. The item and factor GWAS included sex, genotype batch, and the first 10 genomic PCs as covariates. Sex-specific GWAS included genotype batch and the first 10 genomic PCs as covariates.

$h^2_{SNP}$ Z used for item selection for further analysis steps were estimated for the item GWAS in GenomicSEM. These Z estimates were used for item selection criteria. $h^2_{SNP}$ and $r_g$ estimates were run in linkage disequilibrium score regression (LDSC). [7] Liability scale heritability for the items was calculated using the response prevalence in the MoBa sample (with and without genetic data) as the population prevalence and the response prevalence in the subsample with genetic data as the sample prevalence. Genetic correlations were estimated with autism, ADHD, and schizophrenia using summary statistics munged using the sum of effective sample size so a sample prevalence of 0.5 was used. Population prevalence used were 0.02 for autism, 0.087 for ADHD, and 0.01 for schizophrenia.

*Multiple Testing Corrections*

Although the study was exploratory in nature, multiple testing corrections were performed for several of the analytic steps for reference. For the outcome models in the phenotypic data, , adjusted p-values were calculate using FDR method adjusting for 353 tests (16 -outcomes * 11-factors * 2-sex) in the univariate models and 32 in the multivariate (16-outcome models * 2-sex). For the estimated genetic correlations of the items and factors with the PGC GWAS the FDR method was also used to calculate adjust p-values based on 138 tests ((11 -factors + 35 items) * 3 -PGC traits). These were calculated using 4.1.2 of the R package *stats.*

### Supplementary Results

*Exploratory and Confirmatory Factor Modeling*

Procedures to determine the optimal number of factors to retain indicated high dimensionally underlying neurodevelopmental traits as parallel analysis identified 13 factors, 15 factors were identified using eigenvalues, the optimal coordinates method identified 3, and acceleration factor 1. Because of the range in number of factors identified across the different methods we ran 15 EFA models extracting 1-15 factors in one half of the sample. Based on fit indices (CFI, TLI, SRMR, RMSEA, Supplementary Table S5) and interpretability of the identified factors we chose to only run models based on the 9,10, and 11 factor in the other half of the sample as confirmatory factor models. Models with fewer factors in the EFA contained factors that were theoretically hard to interpret or left all items in certain domains of neurodevelopment (i.e., motor development) not well represented by a factor. Models with fewer factors also did not "collapse" the smaller more specific factors from the 9-11 factor models into larger factors in a theoretically interpretable way. Models with more factors in the EFA contained factors that had less than 3 items loading onto them or factors hard to theoretically interpret. Factor loading estimates from the 1-15 factor EFA are presented in Supplementary Tables S6-19.

Besides the 9,10, and 11-factor models based on the results of the EFA, a sensitivity analysis allowing the language factor to correlate with the residual variance of the item measuring excessive talking (an indicator of the impulsivity factor) was also run. This showed that this item was driving the slight negative correlation between impulsivity and language factors but had minimal improvement to the overall fit of the model. A sensitivity analysis only including one sibling from each family in MoBa improved the fit of the 11-factor model in the half sample. Fits for these analyses are in Supplementary Table S20. Three items were removed from the 11-factor model for the CFA step because they did have loadings over 0.4 onto any factor in the EFA. These were items *"poorly coordinated or clumsy"* from the CBCL, *"have any particular friends or a best friend"* from the SCQ, and a MoBa specific question "*high pain threshold".* In the CFA step, a few poorly endorsed items had estimated loadings slightly over 1 in the 11 factor model, constraining these values to be below one did not lead to a significant decrease in model fit.

The factors for the 11-factor model include five factors covering communication and social behaviors. The factors nonverbal communication and joint attention, language and verbal communication, social attention and interest, and play had indicators from the SCQ, M-CHAT, ESAT, and/or ASQ language subscale all of which were designed to screen for autistic traits or language development. The other factor, Prosocial behaviour, had indicators entirely from the SDQ prosocial scale as well as one item asked alongside the SDQ scale in MoBa. The factors corresponding to ADHD traits (waiting, inattention and overactivity, and impulsivity) were made up of the CBCL and MoBa specific questions derived from DSM criteria for ADHD. Indicators for the two repetitive behavior factors (repetitive and restricted behaviors and interests (RepBehavior), repetitive and idiosyncratic speech) were made up of SCQ items and, for RepBehavior, one additional M-CHAT item.

*Common genetic variance underlying early neurodevelopmental traits*

Three of these SNPs were associated with multiple factors (rs61775569, rs12967622, rs10956955 in LD with rs4961212, Table S36). three genes associated with specific factors ($p < 2.682 \times 10^{-6}$; Tables S37-47). The motor factor was associated with *CNGB3* ($p = 1.53 \times 10^{-6}$), while the prosocial behavior factor was associated with *RSRC1* ($p = 3.95 \times 10^{-7}$) and *ADAMTS17* ($p = 8.19 \times 10^{-7}$).

*Genomic structure modeling and specificity of SNP effects*

The smoothed estimated genetic correlation matrix of all chromosomes of the selected item GWAS is presented in the main text (Figure 5). Besides the main cluster of the prosocial behavior items the other two possible clusters based on the correlation matrix were made up of items covering ADHD traits, repetitive and restricted behaviors and interests, and play behaviors and were thus less interpretable. An EFA was run using this correlation matrix. The traditional eigenvalues method indicated eight factors to be extracted in an EFA at the genomic level and thus 1-8 factors were extracted.

The motor, prosocial behavior, RepBehavior, and inattention factors were recreated via a CFA at the genomic level. Only the prosocial behavior factor demonstrated an exceptional fit (CFI = 1, SRMR = 0.095; Supplementary Figure S14) and exhibited strong and significant loadings for most items. The inattention and overactivity factor had significant loadings for all items but as it was comprised of three items, fit indices could not be estimated. Several of the items in the inattention and overactivity factor also had low $h^2_{SNP}$ Z on the edge of our threshold. The repetitive and restricted behaviors and interests had excellent fit (CFI = 0.99, SRMR = 0.083) but non-significant loadings for all items. The motor factor did not converge.

We only performed a subsequent common factor GWAS for the prosocial behavior factor, which did not yield any genome-wide significant loci but identified more Qsnp than SNP hits 6 independent SNPs hits and 17 $Q_{snp}$ at a suggestive association threshold ($p < 5 \times 10^{-5}$; Supplementary Tables S57-58). Removing the item that did not significantly load onto the common genetic factor, the common factor GWAS identified 7 SNPs and 9 $Q_{snp}$ at the same threshold (Supplementary Figure S15; Supplementary Tables S59-60).

### Supplementary Figures

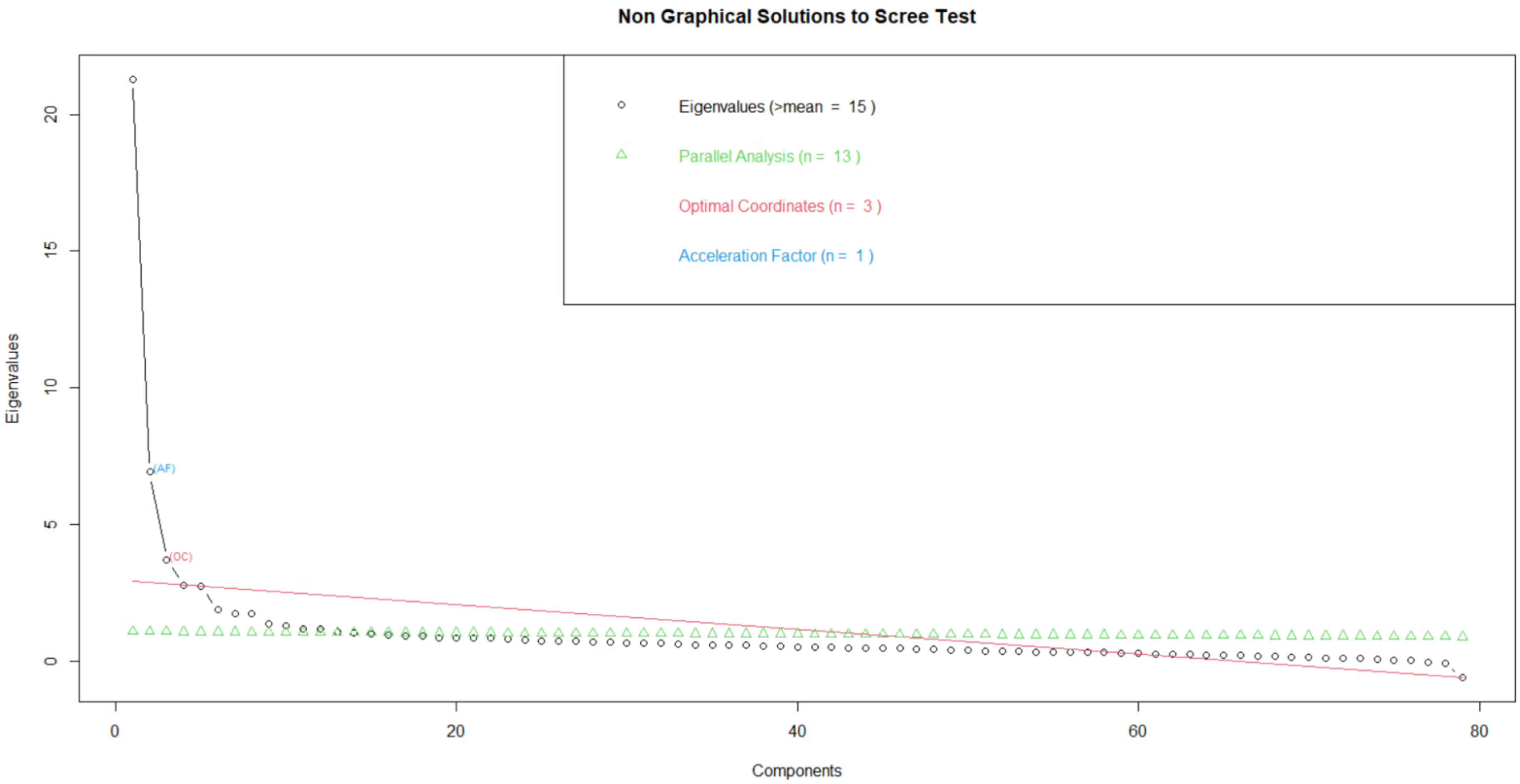

*Figure S1: Scree plot for exploratory factor analysis using 79 items. Results of parallel analysis, acceleration factor, and optimal coordinates presented.*

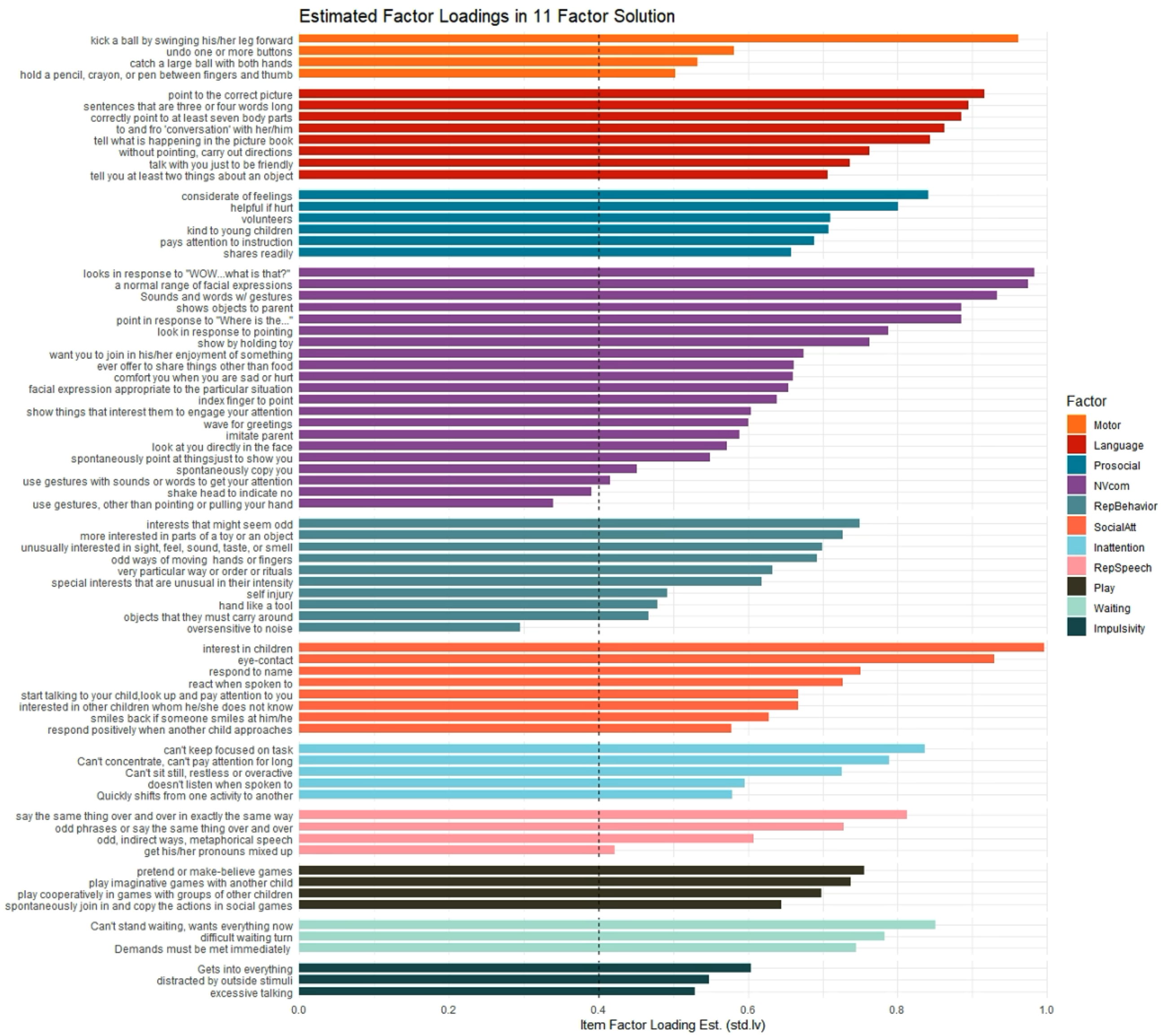

*Figure S2: Estimated standardized factor loadings from the 11-factor correlated factor model run in the full sample. Dotted line denotes a 0.4 factor loading.*

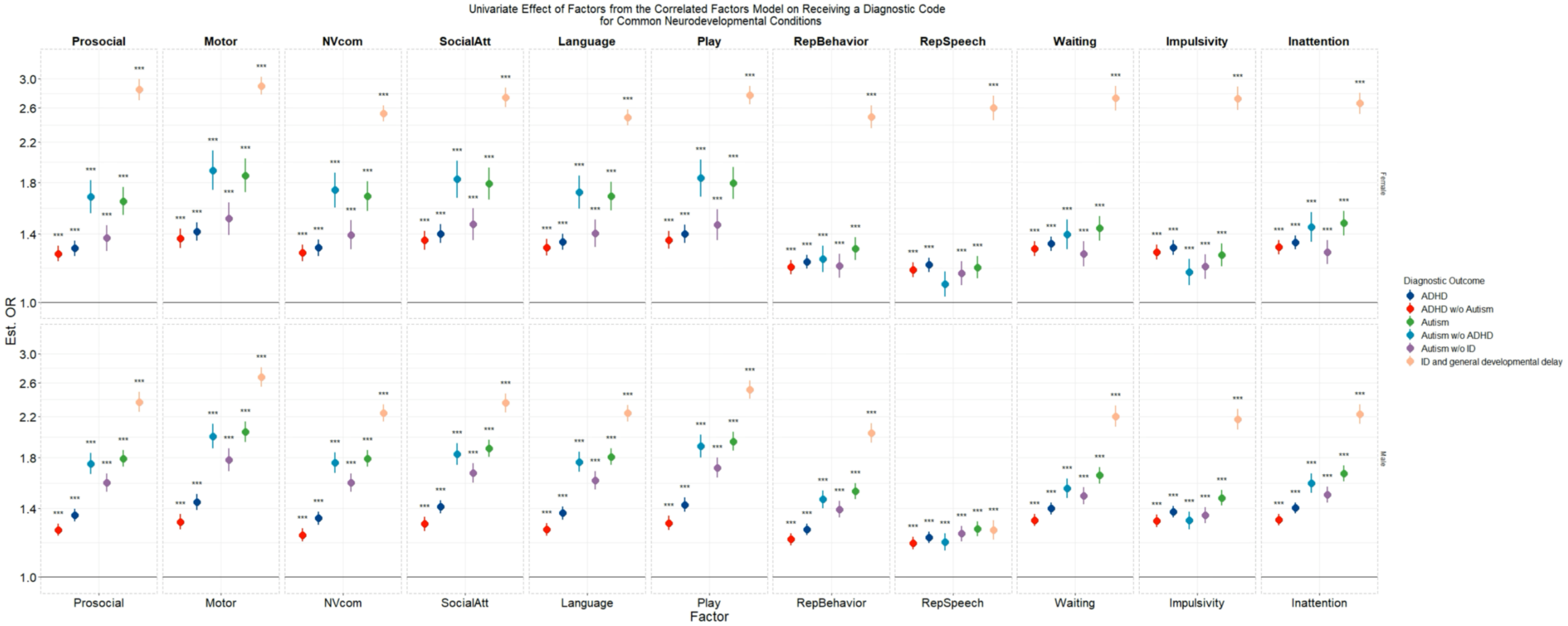

*Figure S3: Estimated effects of the factors independently effecting the main diagnostic outcomes (ADHD, Autism, and Intellectual Disability/General Developmental delay). Effects are presented as odds ratios calculated from the exponential of the standardized beta value from the logistic regression in the measurement models. 95% CI are presented. "*", "**", "***" denote p <0.05, <0.01, and <0.001 respectively, after multiple testing correction.*

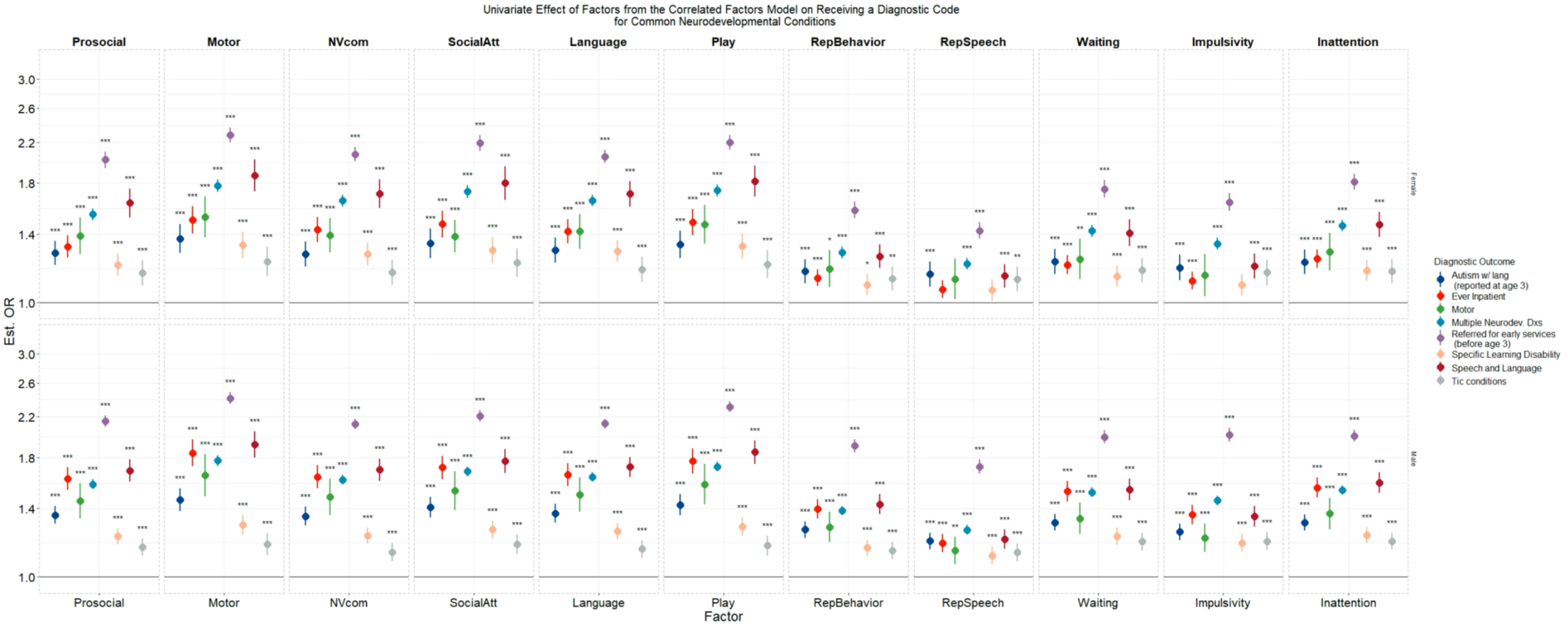

*Figure S4: Estimated effects of the factors independently effecting the additional diagnostic outcomes. Effects are presented as odds ratios calculated from the exponential of the standardized beta value from the logistic regression in the measurement models. 95% CI are presented. "*", "**", "***" denote p <0.05, <0.01, and <0.001 respectively, after multiple testing correction.*

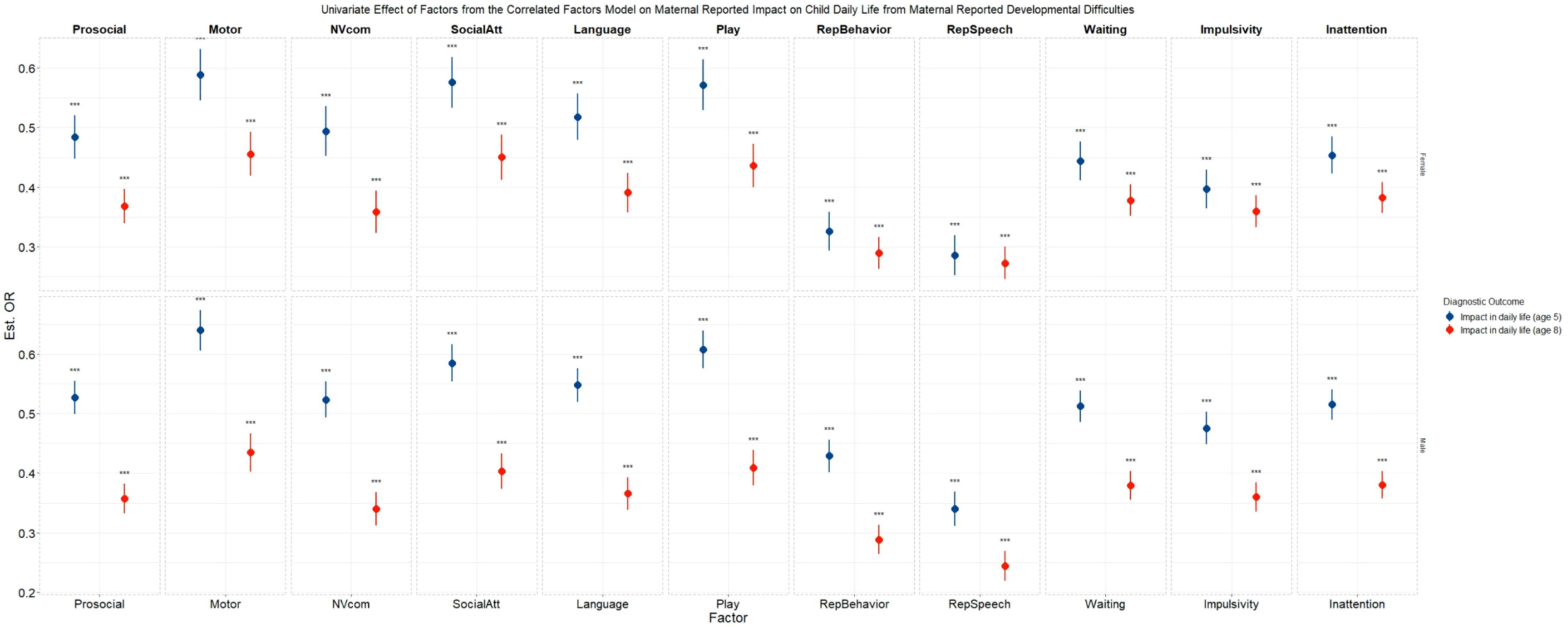

*Figure S5: Estimated effects of the factors independently effecting maternal reported impact in daily life due to developmental difficulties.* 95% CI are presented. *Effects are std. beta values. "*", "**", "***" denote p <0.05, <0.01, and <0.001 respectively, after multiple testing correction.*

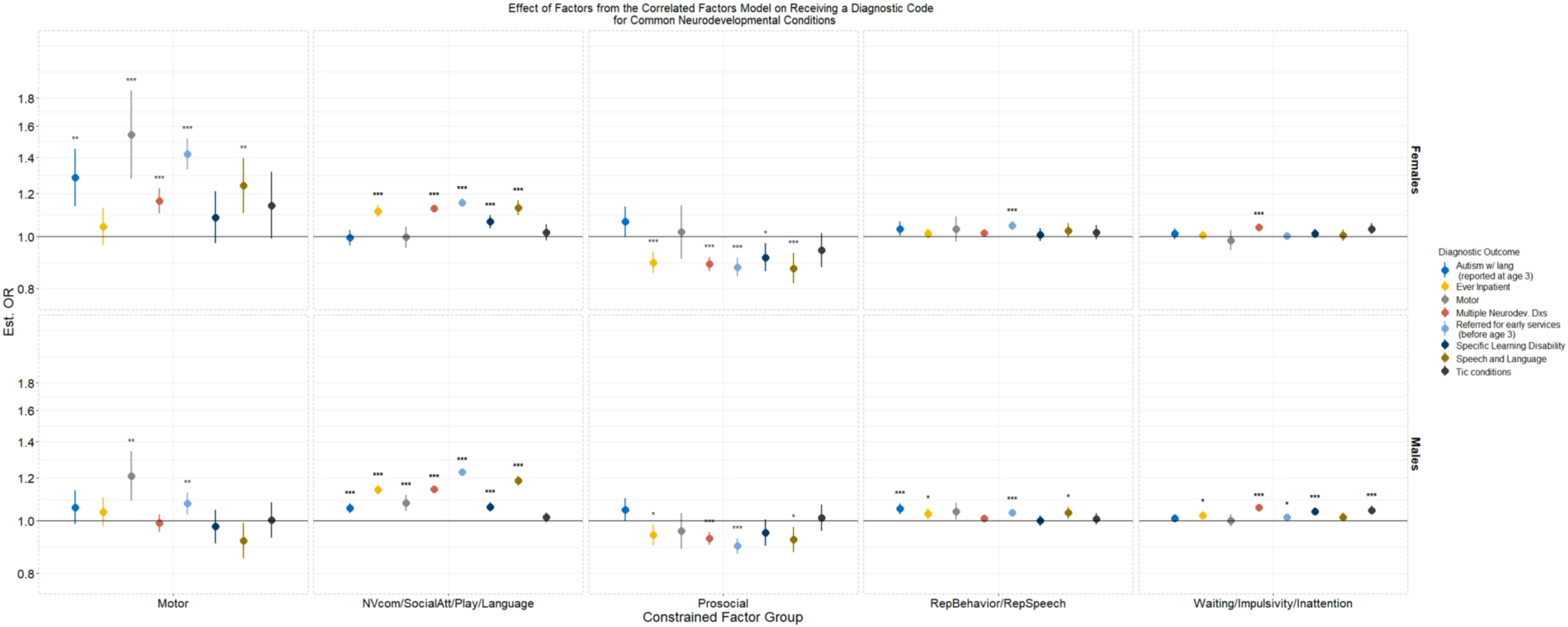

*Figure S6: Estimated effects of factors from the correlated factor model in a multivariate regression controlling for the effects of all factors for the additional diagnostic outcomes not presented in the main results. Effects are presented as odds ratios calculated from the exponential of the standardized beta value from the logistic regression in the measurement models. 95% CI are presented. Due to high correlations amongst domains in the broad areas of social communication (the language & verbal communication, nonverbal communication and joint attention, play, and social attention and interest factors), ADHD-associated traits (the inattention and overactivity, waiting, impulsivity factors), and repetitive and restricted behaviors (the repetitive and idiosyncratic speech and repetitive and restricted behaviors and interests factors) effects of these factors were constrained to be equal to avoid collinearity issues.''*'', "**", "***" denote p <0.05,<0.01,and <0.001 respectively, after multiple testing correction*

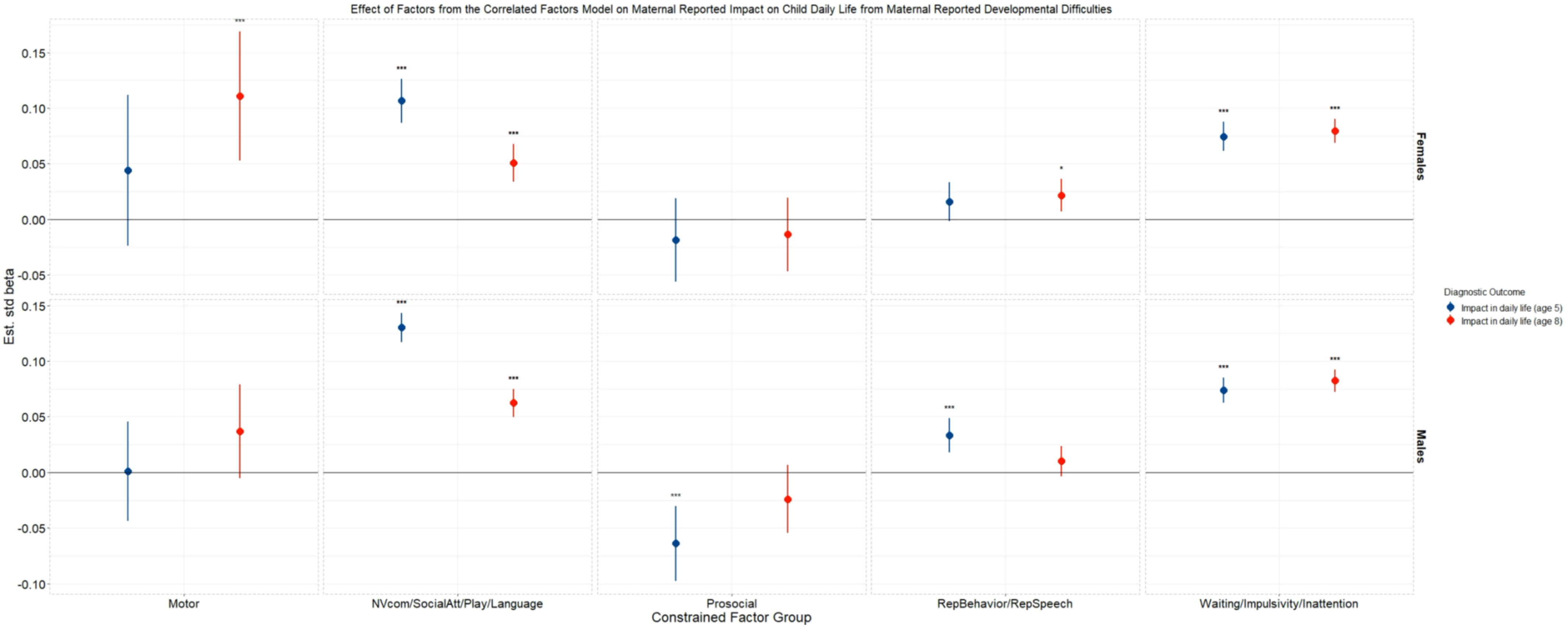

*Figure S7: Estimated effects of factors from the correlated factors model in a multivariate regression controlling for the effects of all factors for effecting maternally reported impact in daily life due to developmental difficulties. Effects are reported as std. beta values. 95% CI are presented. "*", "**", "***" denote p <0.05, <0.01, and <0.001 respectively, after multiple testing correction.*

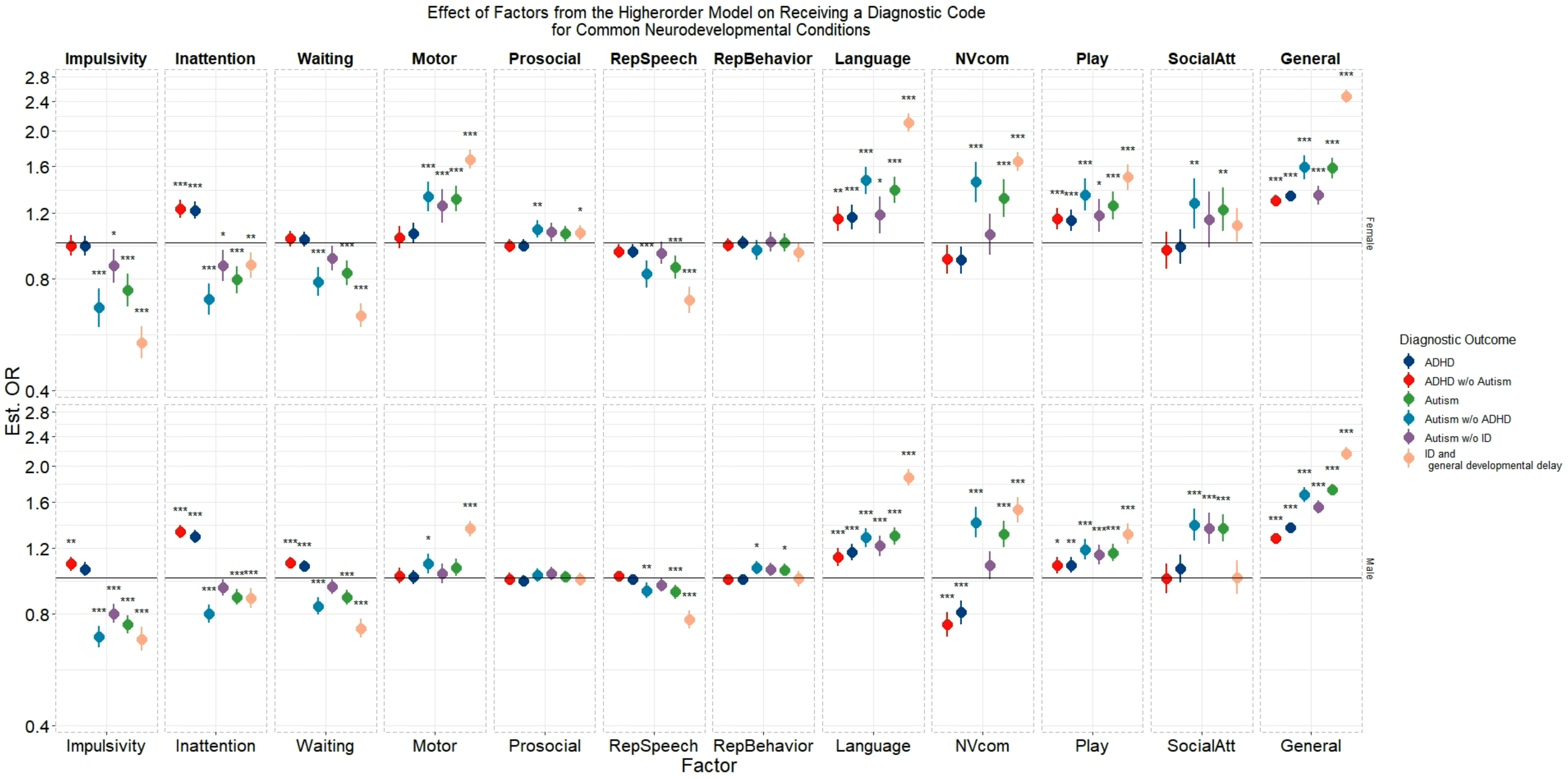

*Figure S8: Estimated effects of the factors from the higher order model on the main diagnostic outcomes. Effects of the specific factors and general factor were estimated in separate models. Specific factors were specified to simultaneously affect the outcome. Effects are presented as odds ratios calculated from the exponential of the standardized beta value from the logistic regression in the measurement models. 95% CI are presented. "*", "**", "***" denote p <0.05, <0.01, and <0.001 respectively, after multiple testing correction.*

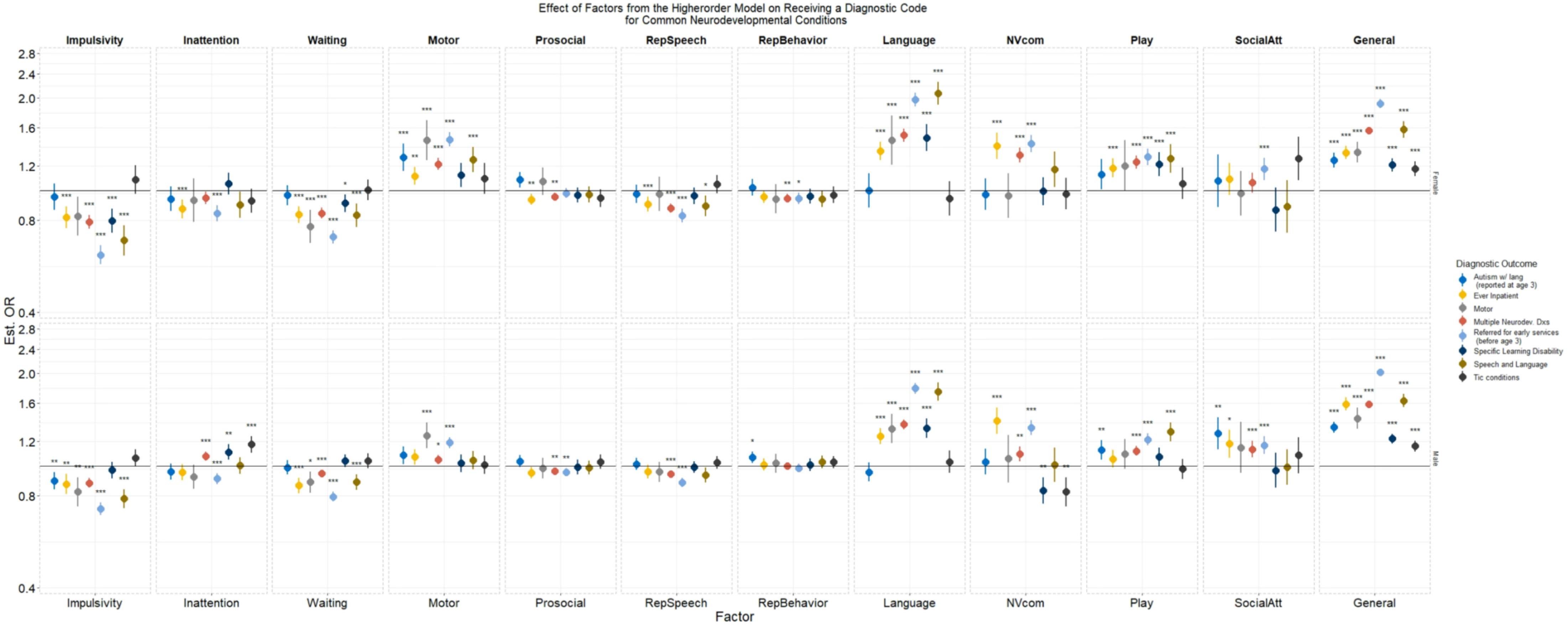

*Figure S9: Estimated effects of the factors from the higher order model on the additional diagnostic outcomes. Effects of the specific factors and general factor were estimated in separate models. Specific factors were specified to simultaneously affect the outcome. Effects are presented as odds ratios calculated from the exponential of the standardized beta value from the logistic regression in the measurement models. 95% CI are presented. "*", "**", "***" denote p <0.05, <0.01, and <0.001 respectively, after multiple testing correction.*

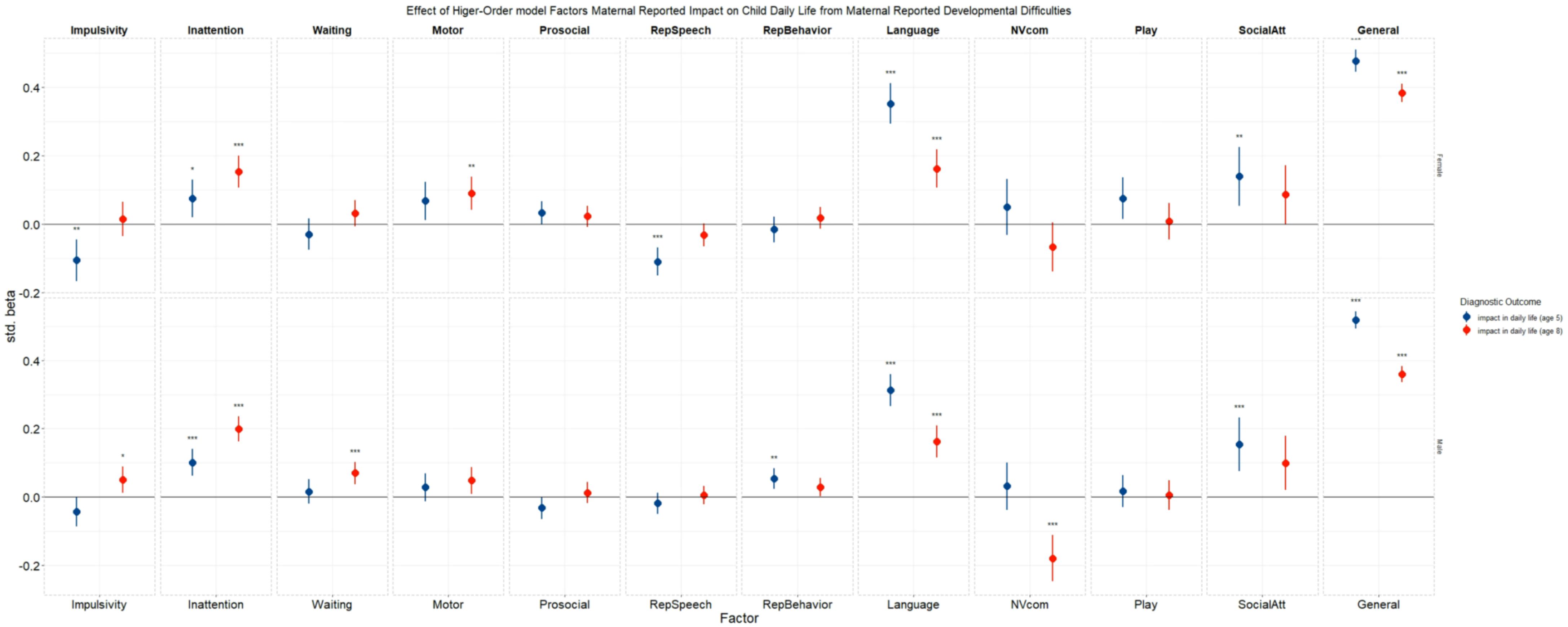

*Figure S10: Estimated effects of factors from the higher order model on maternally reported impact in daily life due to developmental difficulties. Effects of the specific factors and general factor were estimated in separate models. Specific factors were specified to simultaneously affect the outcome. Effects are reported as std. beta values. 95% CI are presented. "*", "**", "***" denote p <0.05, <0.01, and <0.001 respectively, after multiple testing correction.*

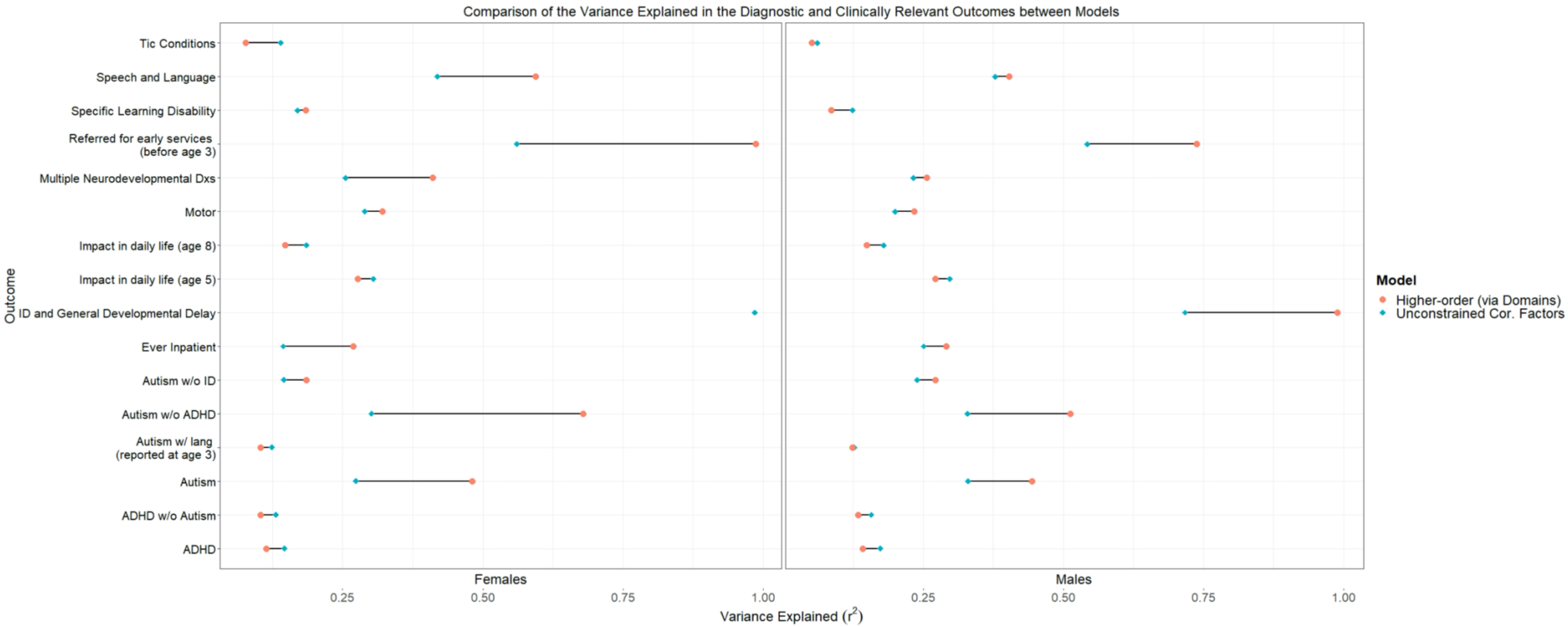

*Figure S11: Estimated $r^2$ of the outcome explained by the specific factors in a higher-order model where the factors moderate the effect of a general factor on the outcome (denoted by an orange circle) vs the factors from a correlated factor model with unconstrained effects (denoted by a blue diamond). $r^2$ for intellectual disability and general developmental delay in girls for the higher order model was estimated significantly over 1 and therefore not presented.*

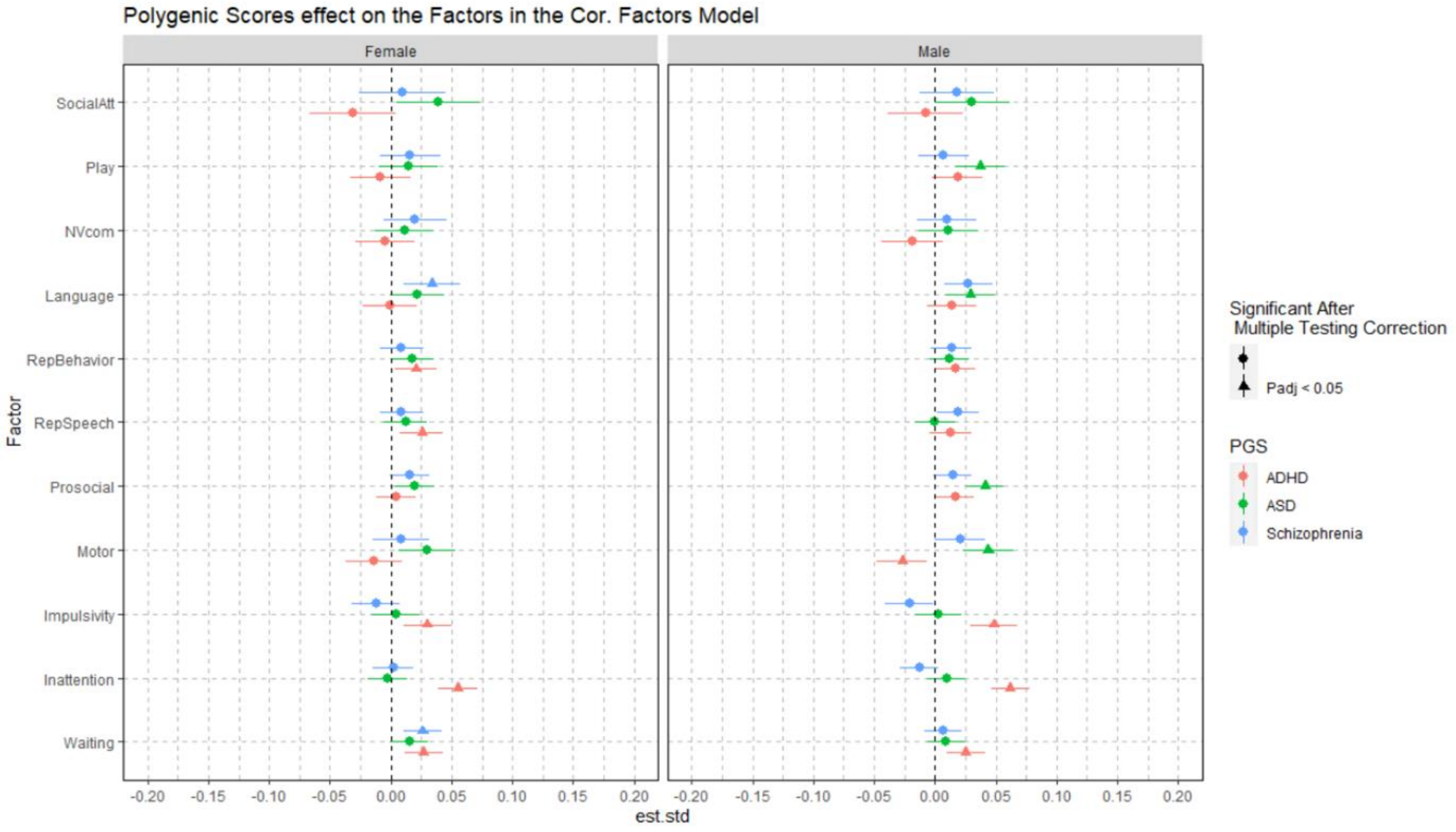

*Figure S12: Effect estimates, presented as std. beta values, of the PGS on the factors from the correlated factor model. 95% CI are presented. PGS were specified to affect the factors simultaneously. Triangle denotes significant ($p < 0.05$) after multiple testing corrections.*

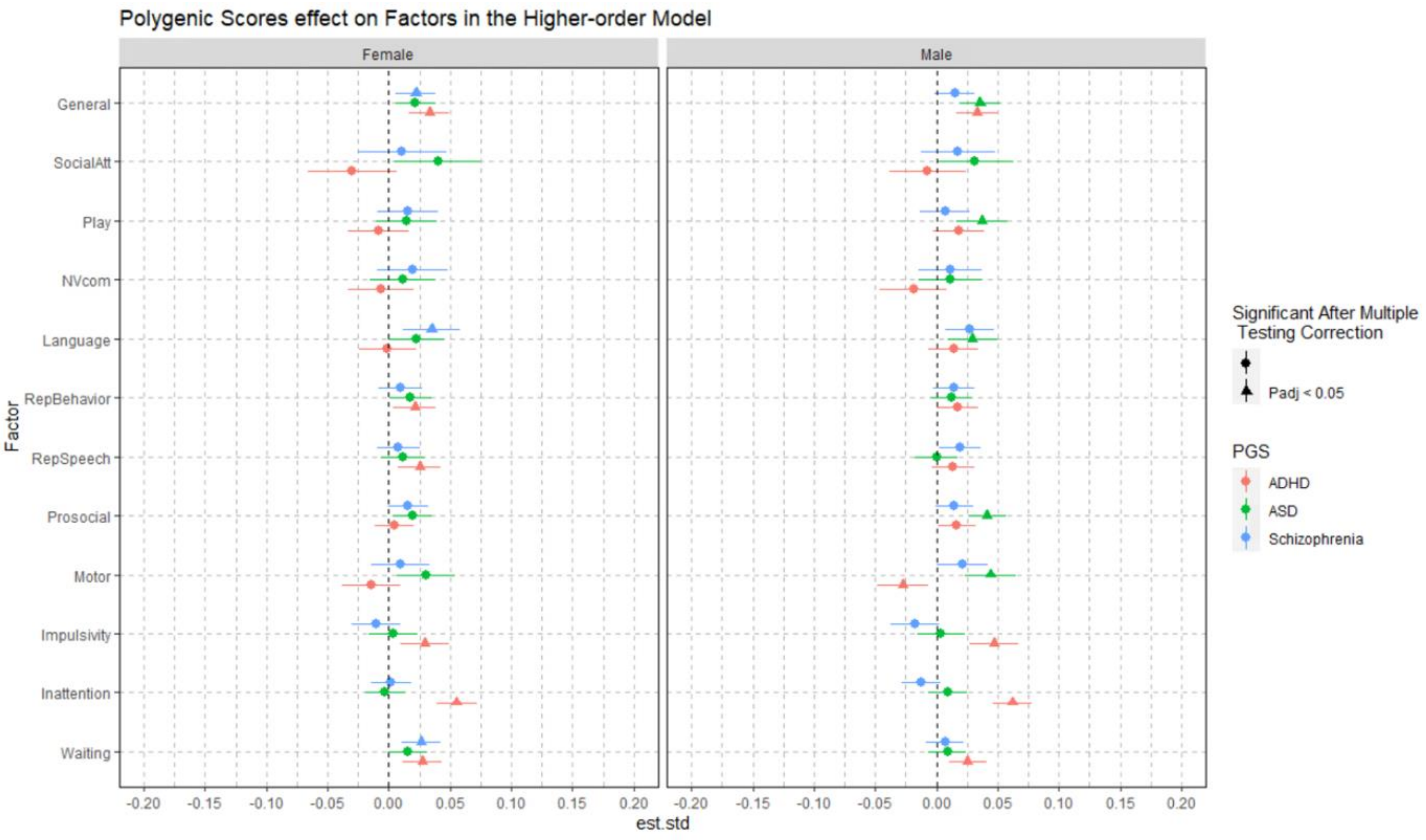

*Figure S13: Effect estimates, presented as std. beta values, of the PGS on the factors from the higher-order model. Effects of the PGS on the specific factors and general factor were estimated in separate models. PGS were specified to simultaneously affect the specific factors. 95% CI are presented. Triangle denotes significant ($p < 0.05$) after multiple testing corrections.*

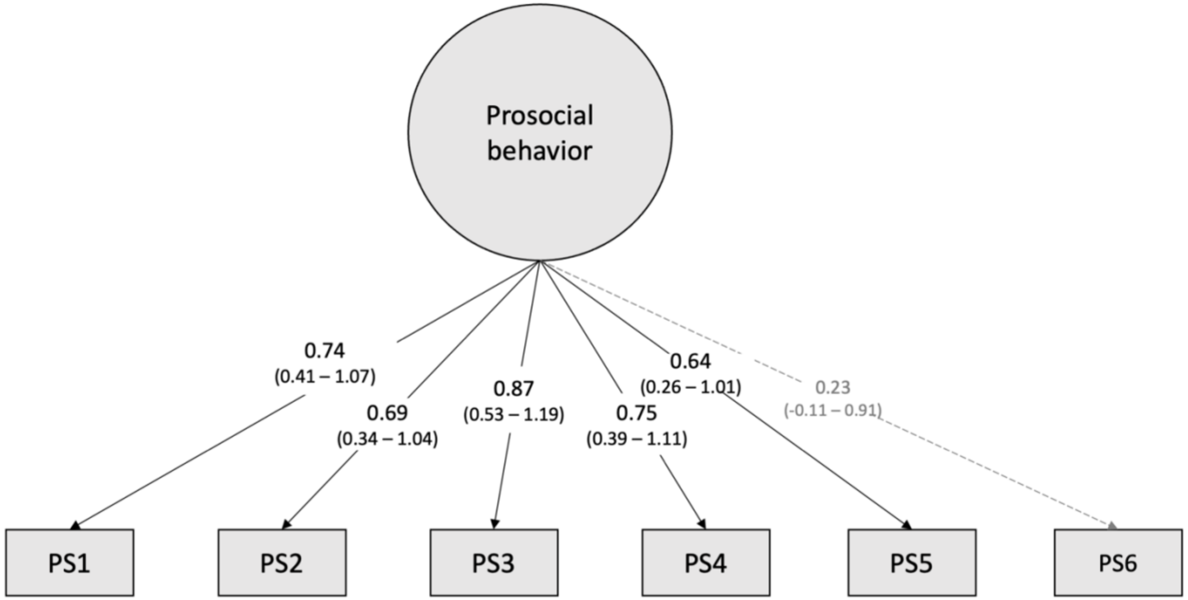

*Figure S14: Estimated factor loadings of the common genetic factor for prosocial behavior. PS1 is item "Your child shares readily with other children, for example treats, toys, pencils", PS2 is item "Your child is helpful if someone is hurt, upset or feeling ill", PS3 is item "Your child is considerate of other people's feelings", PS4 is item "Your child is kind to younger children", PS5 is "Your child often volunteers to help others (parents, teachers, other children)", PS6 is "Your child pays careful attention when you try to teach him/her something new"*

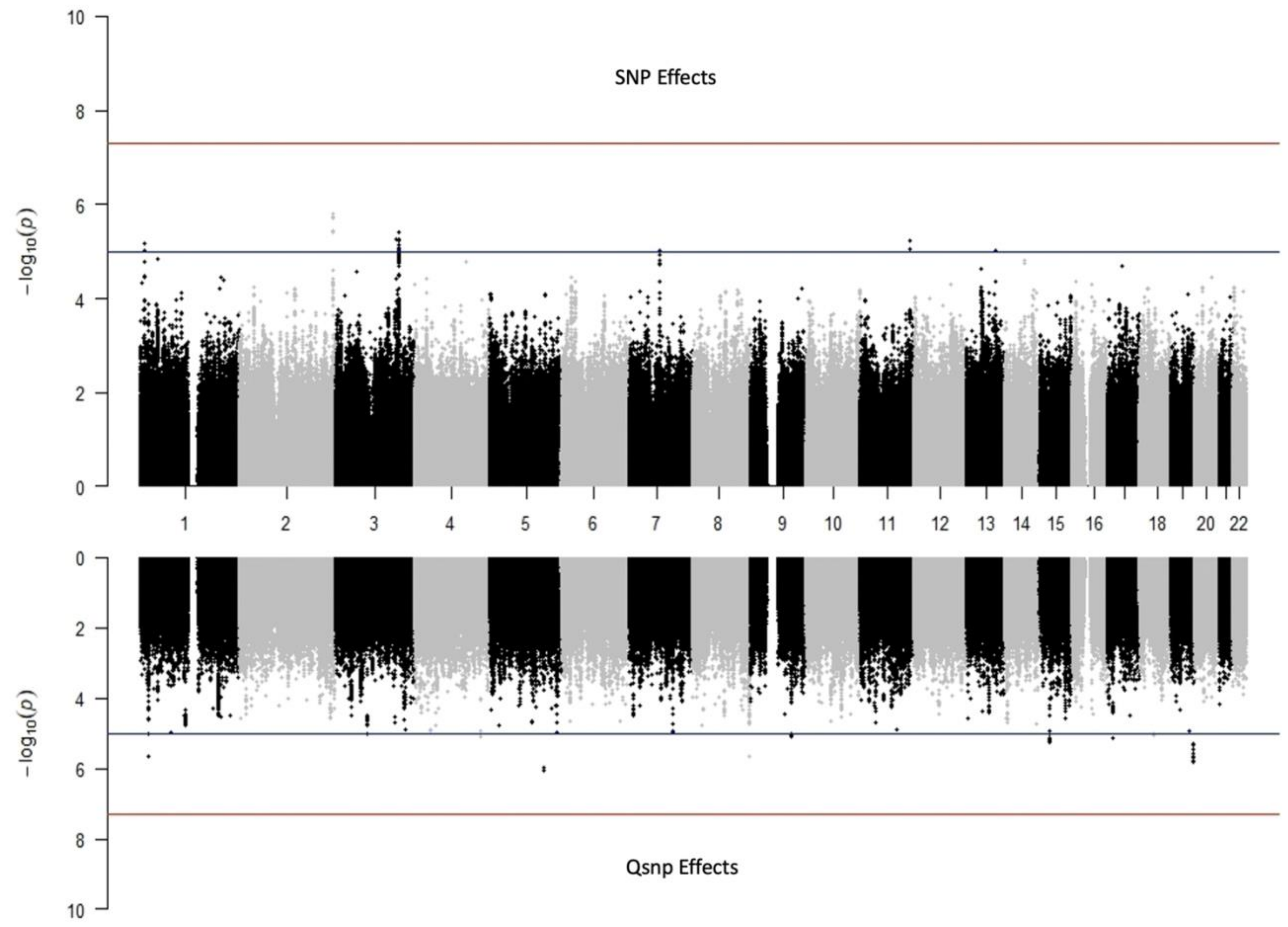

*Figure S15: Miami plot of the common factor GWAS conducted in GenomicSEM of the prosocial behavior factor with only the significantly loading items included. The top plot displays SNP effects and the bottom Qsnp effects. Red line denotes genome-wide significance (p<5×10-8) while the blue denotes suggestive association threshold of (p < 5 x10-5)*
